# High-frequency oscillations and interictal epileptiform discharges predict infantile spasms

**DOI:** 10.64898/2026.07.03.26357202

**Authors:** Sini Hautala, Sinikka La Grassa, Leena Lauronen, Maria Peltola, Maarit Palomäki, Liisa Metsähonkala, Marjo Metsäranta, Henna Jonsson, Eija Gaily, Milja Harju, Mohammad Al-Sa’d, Kirsi Mikkonen, Päivi Nevalainen

## Abstract

Early acquired brain injury is a major risk factor for infantile epileptic spasms syndrome (IESS), which may impair cognitive development, especially if diagnosis and treatment are delayed. However, individual-level prediction of which infants will develop IESS is currently not possible. We assessed whether high-frequency oscillations (HFOs) in scalp EEG or recurrent interictal epileptiform discharges (IED) during the first months of life could predict forthcoming IESS.

Our population-based cohort included 36 infants with cortical injury due to infarction, haemorrhage, infection or trauma involving a large cortical area (≥ anterior/posterior cerebral artery territory or ≥ half of the middle cerebral artery territory), or hypoxic-ischaemic encephalopathy with cortical and deep grey matter involvement. The infants underwent repeated EEGs during the first year of life until 12 months of age or until IESS diagnosis. HFOs during sleep were scored both visually and automatically, whereas IEDs were assessed visually only. We tested whether HFO rate increased during the first year of life using a mixed-effects model with within- and between-subject random effects. Using only EEGs recorded prior to IESS diagnosis, we evaluated whether HFO rate or recurrent IEDs could predict IESS development by training a ridge-regularized logistic regression model with exhaustive leave-2-subjects-out cross-validation.

Eleven infants (31%) developed IESS. HFO rate increased with age in both groups but more steeply in the IESS group [within person slope *β* = 2.43 (IESS) vs. 0.06 (no-IESS) units/month, *P* < 0.001]. The logistic regression model showed that both HFO rate [AUC 0.801 (95% CI 0.668, 0.936)] and recurrent IEDs [AUC 0.826 (95% CI 0.720, 0.932)] were able to predict forthcoming IESS. However, in a multivariable model, only recurrent IEDs remained independently associated with IESS, and HFO rate did not add predictive value.

The marked increase in HFO rate toward IESS diagnosis supports their role as a biomarker of epileptogenesis. During the first months of life, HFOs and recurrent IEDs performed equally well in predicting subsequent IESS. However, IEDs are easier to apply to clinical practice.

## Introduction

An acquired early brain injury due to perinatal stroke,^1^ hypoxic ischemic encephalopathy (HIE),^2^ complications of prematurity^3^ or CNS infection^4^ constitute a major risk factor for developing epilepsy. Seizures often begin in infancy, typically presenting as infantile epileptic spasms syndrome (IESS).^5^ This severe epileptic encephalopathy may adversely affect cognitive development, especially if the diagnosis and, consequently, treatment is delayed.^6^ Therefore, the identification of infants at the highest risk for IESS is crucial.

Despite many identified risk factors,^7–10^ there are no established protocols for individual-level prediction of IESS, particularly in other aetiologies than HIE. Furthermore, there is limited data on how EEG evolves towards hypsarrhythmia (the pathognomonic EEG pattern of IESS), and which early electrophysiological phenomena would be indicative of the ongoing process leading to the onset of spasms.^9,11–13^ An early identification of infants who later develop IESS would enable more personalized follow-up plans, implementation of potential preventive therapy, as in the case of vigabatrin in tuberous sclerosis,^14^ and earliest possible treatment at epilepsy onset.

High-frequency oscillations (HFOs) known as ripples (80–250 Hz) and fast ripples (250–500 Hz) are promising EEG biomarkers of epilepsy, and there is growing evidence on possible applications of HFOs also in IESS. During neonatal period, interictal HFOs are more frequently observed in neonates later developing IESS.^15^ During IESS, ictal fast oscillations (40–150 Hz) are observed in high numbers,^16^ and increased HFO rates and coupling between HFOs and slow-wave activity associate with active spasms.^17^ Once the treatment of IESS is initiated, the number of HFOs decreases,^16,18^ and preliminary evidence suggests that post-treatment HFOs can distinguish the non-relapse group from those who later relapse.^19^

Little is, however, known about the occurrence of HFOs prior to IESS onset.^15^ Rodent models have demonstrated an association between recurrent spontaneous seizures and either HFO appearance, HFO rate or both after induced status epilepticus^20^ and traumatic brain injury,^21–23^ thereby suggesting that HFOs could either play a role in epileptogenesis or, at least, be indicative of it. To date, no longitudinal cohort studies on HFOs in infancy exist.

We set out to investigate the evolution and predictive value of HFOs during the first year of life in infants with a known high risk for IESS due to early brain injury. We hypothesized that in infants developing IESS, HFOs appear already during the first few months of life and HFO rates increase in the period leading up to the IESS diagnosis. Furthermore, we hypothesized that HFO occurrence, HFO rate or both would differentiate infants developing IESS from those who do not. As a secondary analysis, we assessed whether analysing HFOs adds to the predictive value of standard visual EEG analysis based on interictal epileptiform discharges (IEDs).^9,12^

## Materials and methods

### Patients

Our population-based cohort included 36 infants born in the Hospital District of Helsinki and Uusimaa (HUS) between years 2019 and 2023 and who were, before the age of 1 year, diagnosed with a large brain injury known to pose a significant risk (at least 25%–30%) for IESS (Fig. 1A).^7,9,24,25^ The inclusion criteria were either (i) a cortical injury due to infarction, haemorrhage, infection or trauma encompassing an area at least equivalent to the entire territory of the anterior or posterior cerebral artery or approximately half of the middle cerebral artery territory in visual MRI analysis; (ii) HIE grade II–III with injury of both the cortex and the basal ganglia in MRI (MRI criteria 2B or 3)^26^; (iii) grade IV intraventricular haemorrhage (including haemorrhagic infarction within the parenchyma); or (iv) grade III–IV periventricular leukomalacia (PVL) involving porencephalic cysts. A clinically indicated brain MRI was mandatory for enrolment. Altogether 27 of the included infants took part in an ongoing national, multicentre randomised controlled trial of treatment practices aiming to predict and prevent IESS (PREV-IS).

**Figure 1.**
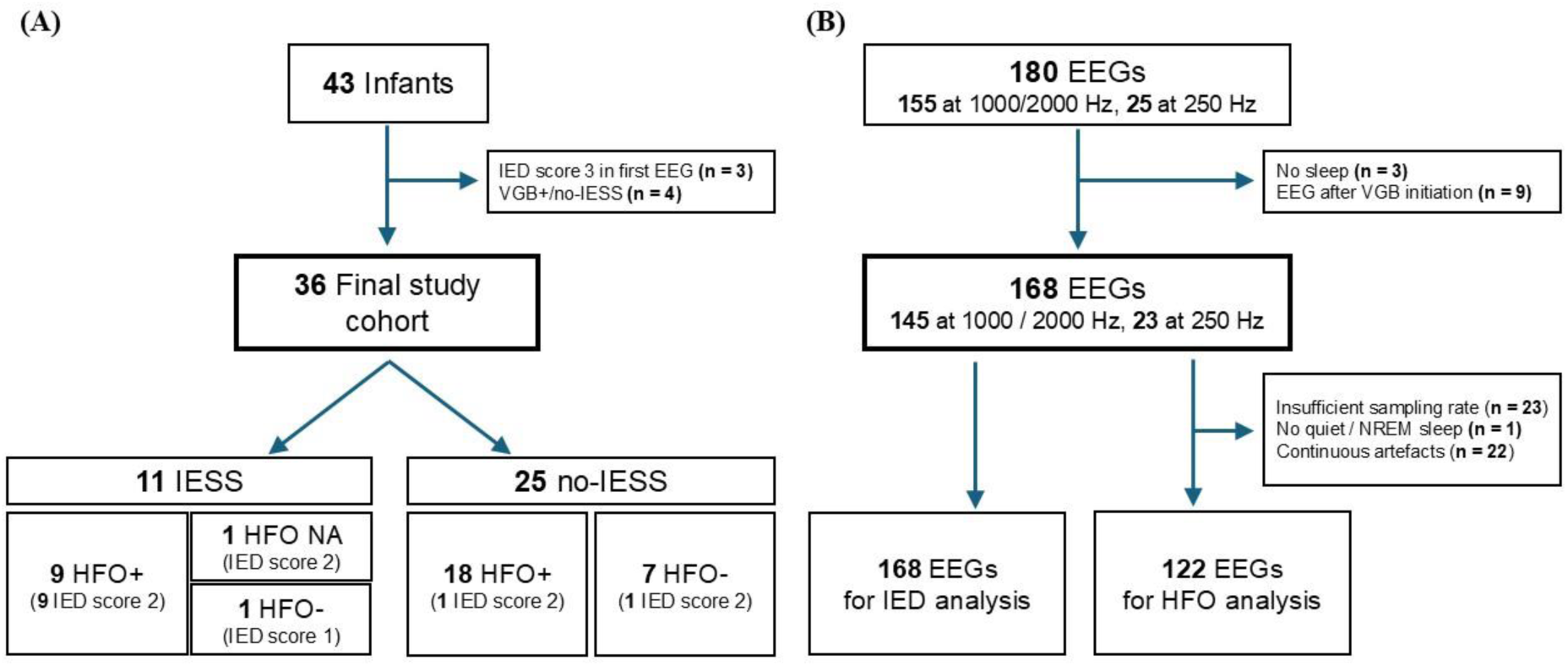
Patient and EEG flow charts. **(A)** Patient flow chart including the IESS (infantile epileptic spasms syndrome) status, occurrence of HFOs (high frequency oscillations) and fulfilment of IED (interictal epileptiform discharges) criteria (score 2) in the final study cohort. VGB+/no-IESS = treated with vigabatrin without developing IESS. IED score 3 = EEG diagnostic of infantile epileptic spasms syndrome. NA = not analysable. **(B)** EEG flow chart depicting the EEG exclusion criteria at each phase. VGB = vigabatrin; NREM sleep = non-rapid eye movement sleep; IED = interictal epileptiform discharges; HFO = high frequency oscillations.

The exclusion criteria were (i) diagnosed IESSM; (ii) vigabatrin treatment for any reason by the time of the first EEG; or (iii) vigabatrin treatment without the development of IESS during the follow-up (possible preventive effect of vigabatrin on the occurrence of IESS). However, we did include infants who received vigabatrin during the follow-up but eventually developed IESS. From such infants, only EEGs before vigabatrin treatment initiation were included (see Methods / Neurophysiology and Fig. 1B). Acute symptomatic neonatal seizures treated according to standard hospital protocols without vigabatrin did not preclude participation.

### Ethics approval and research permit

The institutional research review board of HUS diagnostic centre approved this retrospective registry study (24-HUS/453/2026) and waived for consent due to its observational nature. The prospective PREV-IS study (in which some of the infants participated) was approved by TUKIJA (nationwide medical ethics committee in Finland) and research review boards (decision numbers 21/06.00.01/2018 and HUS/26/2018).

### Neuroimaging

All infants underwent brain MRI as part of the clinical diagnostic process (and therefore, at variable ages) using 1.5 (Siemens Magnetom Avanto Fit, Siemens Healthcare GmbH, Erlangen, Germany; Siemens Magnetom Aera, Siemens Healthcare GmbH, Erlangen, Germany; Philips Ingenia, Philips Medical Systems, Best, The Netherlands) or 3Tesla (Philips Ingenia, Philips Medical Systems, Best, The Netherlands; Siemens Magnetom Skyra, Siemens Healthcare GmbH, Erlangen, Germany) scanner. The imaging protocol included at least T1-weighted axial, T2-weighted axial and coronal, and diffusion-weighted axial images.

Blinded to the neurophysiological findings and clinical outcome, an experienced neuroradiologist (author MPa) re-evaluated all brain MRIs as to the fulfilment of inclusion criteria. The laterality and type of observed lesions were further categorized according to predefined criteria (for HIE, Shankaran^26^; for stroke, Kirton^27^ and Govaert^28^).

### Neurophysiology

#### EEG recordings

All EEGs (*n* = 180) were recorded in the clinical neurophysiology units of Helsinki University Hospital.

The 27 infants participating in the PREV-IS study underwent frequent EEG monitoring at 1, 2.5, 4, 5.5, 7, 9 and 12 months of age (corrected age in preterm infants) or until the primary end point of the study i.e. the diagnosis of IESS. The infants not included in the PREV-IS - study followed our hospital’s current clinical follow-up protocol (EEG at 3, 5 and 12 months of corrected age). For patients whose qualifying brain injury diagnosis was made only later in infancy, the above-mentioned protocols were followed from the time of inclusion diagnosis onward. Consequently, each infant underwent 1–10 EEGs at varying time points. Infants had additional EEGs if clinically indicated, e.g. due to suspected seizures.

Most EEGs (*n* = 155) were acquired at a sampling rate of either 1000 or 2000 Hz using NicoletOne EEG systems equipped with V44 amplifiers (Cardinal Healthcare/Natus, USA; bandwidth 0.53–500 Hz) or Stratus EEG systems (Kvikna Medical, Iceland) with R40 amplifiers (Lifelines Neuro, USA; bandwidth 0.16–524 Hz). In addition, 25 EEGs were acquired at a lower sampling rate of 250 Hz. All recordings employed a Cz reference and 19-channel EEG caps conforming to the international 10–20 system, using sintered Ag/AgCl electrodes (Waveguard, ANT-Neuro, Germany).

#### EEG analysis

Of the 180 EEGs, three EEGs were excluded from further analysis due to lack of sleep and nine EEGs because they were recorded during vigabatrin treatment (see Fig. 1B for EEG exclusion criteria at each phase).

#### HFO analyses

All technically eligible 145 EEGs (23 excluded due to insufficient sampling rate of 250 Hz) from 36 infants were converted to EDF and then analysed in the BrainQuick software (Micromed S.p.A, Treviso, Italy). One neurophysiologist (author SH) first identified and marked artefacts on each channel and annotated the artefact-free periods for vigilance states or as seizure periods.^29,30^ For HFO assessment, we selected up to 5 minutes of NREM stage 2 or 3 sleep or quiet sleep (the first artefact-free segment starting from a sleep spindle if present) per patient. After preliminary screening, altogether 23 EEGs were further excluded due to lack of quiet/NREM sleep (*n* = 1) or due to continuous artefacts (>80% of the selected 5-min segment) on most channels (>50% of all channels or >50% of the channels on a single hemisphere, *n* = 22). Consequently, 122 EEGs from 36 patients entered the final HFO analysis. Possible seizure periods were not analysed.

Two neurophysiologists (authors SH and PN) blinded to the subsequent clinical outcomes and aetiology, manually annotated HFOs using a bipolar double banana montage (Fp2–F8, F8–T4, T4–T6, T6–O2, Fp1–F7, F7–T3, T3–T5, T5–O1, Fp2–F4, F4–C4, C4–P4, P4–O2, Fp1–F3, F3–C3, C3–P3, P3–O1, Fz–Cz, Cz–Pz). The EEG display was divided vertically: standard EEG filtered at 0.5–70 Hz visualized on the left, while the right side showed the same signal filtered between 80–250 Hz using a finite impulse response filter, and a time scale of 1–1.5 seconds per page. HFOs were defined as distinct events consisting of four or more oscillations that clearly stood out from background activity and were not attributable to artifacts. Each rater also assigned a certainty score (CS) to every EEG: 1 = definite HFOs, 2 = probable HFOs, 3 = uncertain HFOs and 4 = no HFOs. Patients for whom the raters gave scores of 1 or 2 were categorized as having certain HFOs. Those with at least one uncertain rating (CS 3) were considered to have uncertain HFOs. Patients who were scored 4 by either of the two raters were considered HFO negative, and only cases in which both raters detected HFOs (definite, probable or uncertain) were classified as HFO positive and included in further rate calculations.

The HFO rates were calculated per minute for each channel in each patient by dividing the total number of HFOs by the duration of analysed, artifact-free time. For each scorer, an overall ripple rate (ORR) was determined by summing all identified HFOs across the included channels and dividing by the total amount of artifact-free time analysed across those channels. Lastly, average ORRs (AORR) were calculated between the two scorers. AORR calculations included HFOs only from channels where both scorers had detected them. AORR was calculated both for the full dataset and separately for the midline (Fz–Cz, Cz–Pz), and left (Fp1–F7, F7–T3, T3–T5, T5–O1, Fp1–F3, F3–C3, C3–P3, P3–O1) and right (Fp2–F8, F8–T4, T4–T6, T6–O2, Fp2–F4, F4–C4, C4–P4, P4–O2) hemispheres. A detailed description of AORR calculation is available in a previous publication.^15^

HFOs were additionally detected by an automatic detector,^31^ separately for frequency ranges 8–140 Hz and 140–250 Hz (Matlab R2023a). We chose to separate these frequency ranges as many previous studies on HFOs in IESS only analysed HFOs <150 Hz,^16,32^ and non-invasive HFO studies in other epilepsy types have indicated that the detected events are most often at the lower end of the ripple spectrum.^33^ Detections during the manually marked artefact periods were excluded.

#### IED scoring

Two neurophysiologists (authors PN, LL, MPe or SV) scored all eligible EEGs (*n* = 168) according to predefined IED criteria^9,12^ using the clinical software (NicOne or Stratus): Score 1: No significant epileptiform or seizure discharge findings i.e. not fulfilling criteria for score 2 or 3; Score 2: Recurrent epileptiform activity: ≥ three 30-s episodes during sleep with (i) ≥5 epileptiform discharges from at least two foci or (ii) ≥10 epileptiform discharges from one focus, and neither clinical spasms nor EEG fulfilling the criteria for score 3; Score 3: EEG diagnostic of IESS: (i) hypsarrhythmia with or without a typical cluster of infantile spasms or (ii) modified hypsarrhythmia or multiple independent spikes with a cluster of infantile spasms. In cases of disagreement, a third neurophysiologist evaluated the recording, and the decision was made by majority vote. For the infants included in the PREV-IS study, the IED scoring was prospective, whereas for those not included in the PREV-IS study, the scoring was done pseudoprospectively blinded to the clinical outcome.

### Outcome

Within the PREV-IS study, an infant was diagnosed with IESS if two neurophysiologists independently evaluated the EEG trace as diagnostic for IESS (score 3; see Methods / IED scoring). For one infant, the IESS diagnosis was made and medication started by a paediatric epileptologist on clinical basis (clusters of epileptic spasms, EEG confirmation not available due to holiday season).

As to infants not participating in the PREV-IS study, a paediatric neurologist (author SLG) retrospectively reviewed the medical records for the date of the IESS diagnosis.

### Statistics

We performed all statistical analyses using RStudio (version 2022.07.1, R Core Team, 2020). Custom R scripts were employed for data preprocessing and analysis. Data visualization was primarily accomplished using ’ggplot2’.^34^ For level of statistical significance, we chose *P* < 0.05. Data normality was assessed with Kolmogorov-Smirnov test.

To assess interrater agreement between the neurophysiologists and the automatic detector, we calculated intra-class correlation coefficient (ICC) estimates (using a two-way mixed-effects model) for the HFO rates. To compare agreement on the certainty scores we employed Cohen’s Kappa.

To test whether HFO occurrence differed between patients who developed IESS and those who did not, we used age-stratified subgroup analysis and Fisher’s exact test.

To test whether AORR increased over the 1-year follow-up, we used a within- and between-subject random-effects (REWB) mixed-effects model to assess both the group (between-subject) and individual (within-subject) levels [see the Supplementary Material (chapter 1.1) for details].^35^ In this analysis, we primarily considered manually scored HFOs of all certainty levels (certain and uncertain). We assessed the results for HFOs from different brain areas (midline, left and right hemisphere), certain HFOs and automatically detected HFOs as a secondary analysis.

To assess whether AORR or IED score predicted the development of IESS across age, we trained a ridge-regularized logistic regression model (*α* = 0) with cubic splines using the glmnet package in R.^36,37^ This model was chosen to assess the overall risk (instead of time-dependent risk) and avoid overfitting in a limited study population with some missing samples. Internal validation was performed using exhaustive leave-two-subjects-out (L2SO) cross-validation (CV), in which every possible pair of patients served as an independent test set. Because the outcome of the model is the probability of IESS at a specific recording timepoint, each EEG was treated as an independent prediction point; thus, predictions and performance metrics were evaluated at the sample level, while all resampling procedures were performed at the patient-level to avoid data leakage. The model building is rationalized in more detail in the Supplementary Material. Model performance and calibration were estimated from out-of-fold (OOF) predictions aggregated across L2SO iterations at the sample level, in line with TRIPOD guidance.^38^ Receiver operating characteristic (ROC) analysis was used to determine the area under the curve (AUC) with 95% confidence interval and the optimal probability threshold at sample level. To enable clinical use of the classifier in selecting patients for example for possible future preventive treatment, we set the threshold to achieve specificity ≥95% while maximizing sensitivity. We prioritized high specificity to minimize false positives, given the potential adverse effects of antiseizure medication (ASM).^39–41^ Calibration was assessed by the intercept, slope, and Brier score. In case internal validation showed an adequate model performance, a final model was refitted on the full dataset and used to derive the predicted risk surface and the age-dependent AORR cutoff curve. See the Supplementary Material (chapter 1.2) for further details.

## Results

### Patients and MRI

Of the 36 patients included in the final analysis, 13 (36%) were female. The median gestational age was 38+6 weeks (IQR 32+2–39+6). Twelve infants (33%) were born preterm. The most common brain injury type was presumed or confirmed perinatal stroke (31/36, 86%). Altogether 15 infants (42%) had bilateral brain injury. Eventually 11 patients (31%) developed IESS (Fig. 1A).

### HFOs

The median number of EEGs accepted for HFO analysis per patient was 3 (range 1–7), and the total number of EEGs was 122 (Fig. 2). Data from 2010 EEG channels were assessable for HFOs (out of the 2196 recorded channels).

**Figure 2.**
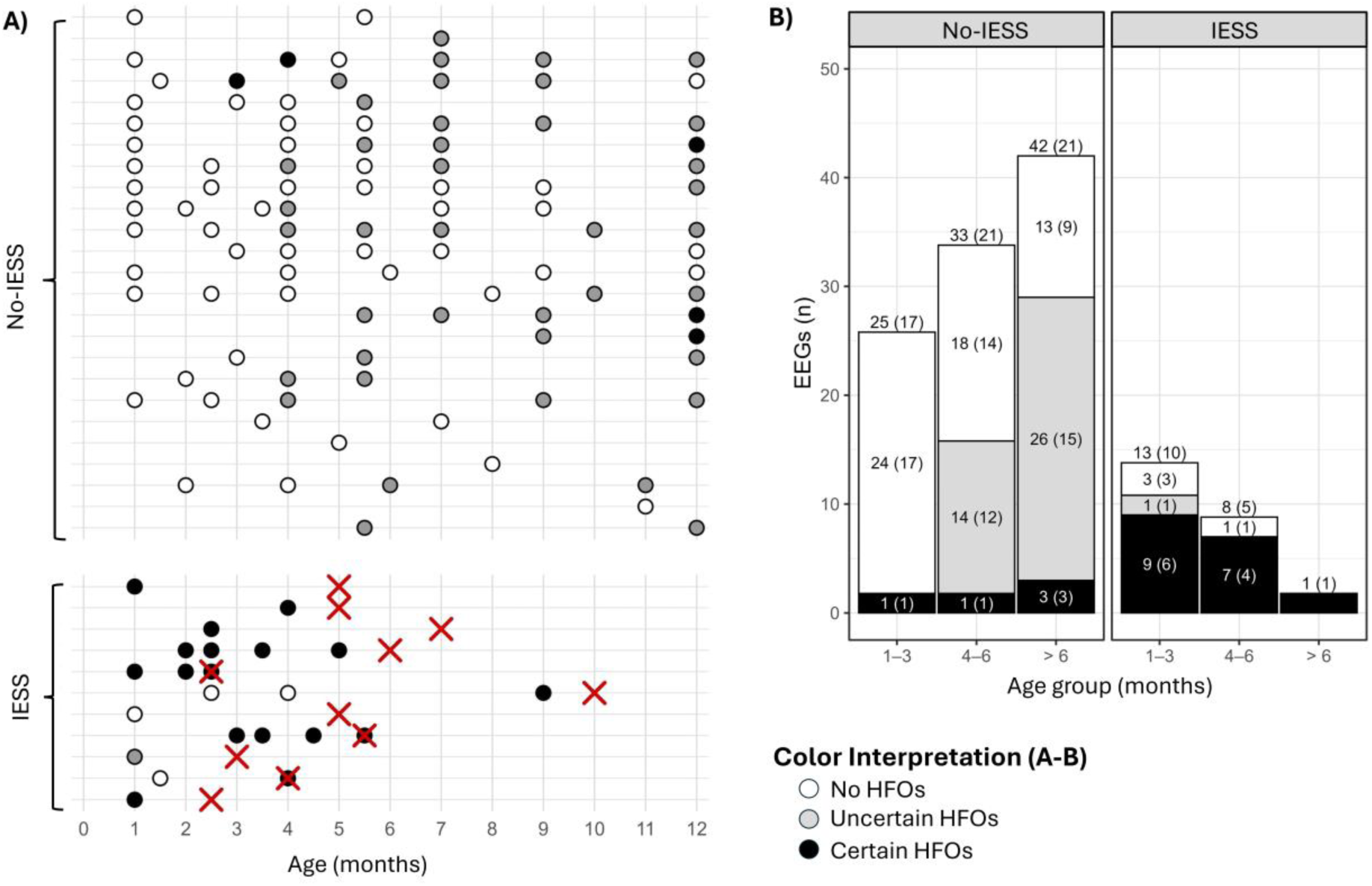
Occurrence of HFOs by IESS (infantile epileptic spasms syndrome) status at patient (A) and group (B) levels. Only EEGs eligible for HFO analysis are presented. HFO certainty is indicated by identical colour coding in both sections A-B. Black = certain HFOs, grey = uncertain HFOs, white = no HFOs. **(A)** Patient-level HFO occurrence and timeline. Patients with and without IESS diagnosis are depicted separately. Each row represents a unique patient. Red X marks the time of IESS diagnosis. For visualization purposes, ages were rounded to the nearest full month. Some infants had additional EEG controls, which is why there are also samples with an accuracy of 0.5 months. The exact ages were used in statistical analyses and treated as a continuous variable. **(B)** Overall HFO occurrences in different age groups. Every EEG analysed for HFOs is depicted. Since most patients had multiple EEGs analysed during the follow-up period, we divided the data into age groups for visualization purposes. Numbers inside the bars indicate the total number of EEGs, with the number of unique patients shown in parentheses.

#### Manually scored HFOs

Both scorers identified HFOs in 63/122 EEGs (51.6%, 22 certain) from 27/36 infants (75.0%, 14 certain). The certainty scores were well correlated between the two raters (overall *κ* = 0.65, *z* = 11.2, *P* < 0.001.; see the Supplementary Material chapter 2.1.1 for details). For the 63 HFO positive EEGs, a total of 1347 channels (out of the 1440 recorded) were analysed, and both scorers identified HFOs on 814 (60.4%) channels. The overall HFO rates were highly correlated between the two scorers [ICC 0.89, *P* < 0.001, 95% CI (0.85, 0.91)]. AORR median was 0.079 (IQR 0.000–0.550; range 0–14.24).

#### Automatic HFO detections

The automatic detection rates in the 80–140 Hz frequency band correlated well with manually scored AORR [ICC 0.73, *P* < 0.001, 95% CI (0.66, 0.80)] with a median rate of 0.63/min (range 0–6.67/min) among all analysed EEGs. On the contrary, in the 140–250 Hz band the automatic rates were consistently higher (median 3.57/min, range 0.36–10.62) and did not correlate with manually scored AORR (ICC -0.119, *P*=0.907) and were, hence, not considered in further analyses (Supplementary Fig. S3).

#### Evolution of HFOs with increasing age in the IESS and no-IESS groups

Among the 1–3-month-old infants, HFOs were rarely present in the no-IESS group, whereas they were present in most infants of the IESS group [OR 58.00 (95% CI 5.47, 3192.06), *P* < 0.001, Fisher’s exact test]. Among the 4–6-month-olds, HFOs were already present in most infants in both groups, and there was no clear association between HFO occurrence and IESS (*P* = 0.182). In the oldest age group (>6-months-olds), there was only one sample in the IESS group preventing statistical testing. (Fig. 2).

To examine how HFO rate evolved with age in children with and without IESS, we used a within-between mixed-effects model. Age was decomposed into within-person and between-person components to differentiate longitudinal changes within the same child from average differences between children of different ages. The model included group (IESS vs. no-IESS) and its interactions with both age components, with random intercepts for participants. The model selection is further described in the Supplementary Material (chapter 1.1). Children with a single measurement (IESS *n* = 6, no-IESS *n* = 4) contributed only to the between-person effect. This method allows true differences in HFO rate between the groups to be separated from apparent differences caused by possible variation in the children’s ages. The within-person coefficient reflects the expected change in AORR for the same child per +1 month. The between-person coefficient reflects how much the AORR differs between children when one child’s average age is one month higher than another child’s.

The model showed that AORR increased with age in both groups, but the growth rate was markedly steeper in the IESS group (Fig. 3). In the no-IESS group, the within-person slope was small but statistically significant [*β* = 0.06 units/month (95% CI 0.01, 0.12), *P* = 0.033]. In the IESS group, the slope was much steeper [*β* = 2.43 units/month (95% CI 2.11, 2.75), *P* < 0.001]. The difference in within-person slopes between groups was significant [+2.37 units/month (95% CI 2.11, 2.75) *P* < 0.001]. Between-patient average age was not significantly associated with AORR levels [between-effect difference +0.32 (95% CI -0.50, 1.13) *P* = 0.438], indicating that group differences were driven primarily by within-person change over time rather than baseline age. Random-effects indicated substantial between-patient heterogeneity (random intercept SD = 1.53, residual SD = 0.97; intraclass correlation coefficient **=** 0.72).

**Figure 3.**
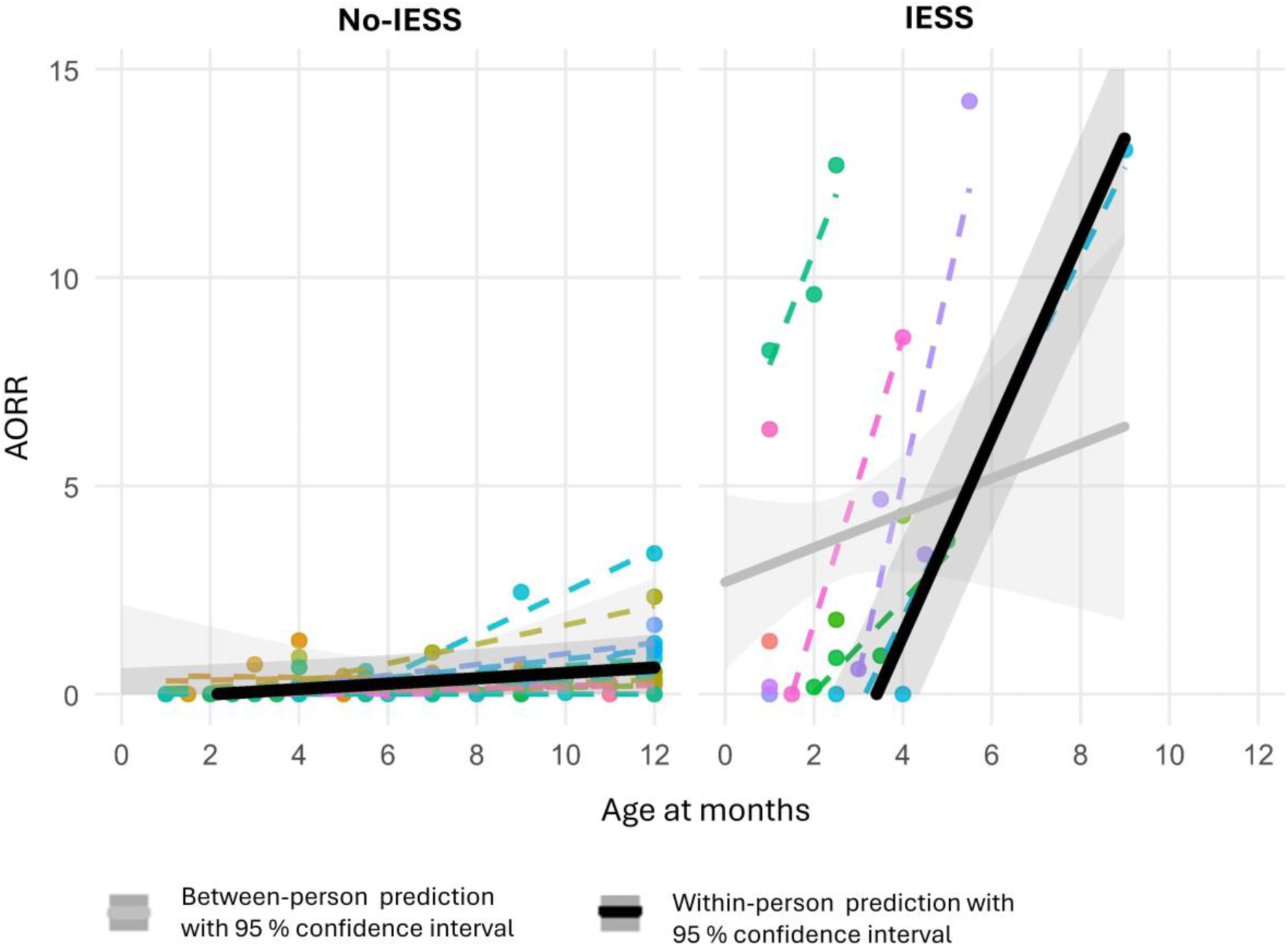
**Mixed-effects model with random intercepts for age and HFO rate** (average overall ripple rate, AORR) **in the no-IESS and IESS** (infantile epileptic spasms syndrome) **groups, together with observed individual AORR trajectories**. Dots show observed AORR values, while dashed coloured lines represent raw individual linear fits (patients with ≥2 measurements). The solid black line represents the population-level fixed-effects prediction at the within-person level with its 95% confidence interval (effect of age change within the same patient). The solid grey line represents the corresponding prediction at the between-person level with 95% confidence interval (effect of differences in average age between patients). In both groups, AORR increases with age within patients, with a markedly steeper increase estimated in the IESS group.

To confirm the robustness of these results, we repeated the above analysis using only certain manually detected HFOs, HFOs originating from defined brain areas and HFOs detected by the automatic detector. Results from analyses using the channel with the maximal HFO rate, the hemisphere with the higher AORR or the midline AORR were concordant with results from the primary analysis. When using only certain HFOs, a statistically significant rate increase over time was observed in the IESS group, but not in the no-IESS group. Automatically detected HFO rates (80–140 Hz) yielded results consistent with those based on manually detected HFOs, but slopes were not as steep. See the Supplementary Material (chapter 2.2) for details.

#### Prognostic value of HFOs in IESS

IESS was diagnosed in 11 infants at a median age of 5 months (range 2.4–9.6), and HFOs were detected in 15 of the 19 (79%, 14 certain) pre-IESS EEGs available for analysis (Fig. 2). Of the 11 IESS patients, nine (82%) had HFOs in at least one of their pre-IESS EEGs (8 certain at some point). The two IESS patients with no HFOs had only a single recording technically suitable for HFO analysis at the age of 1–1.5 months. Altogether 25 patients did not develop IESS and had a total of 100 EEGs available for HFO analysis. HFOs were detected in 45 EEGs (45.0%, 5 certain) from 18 different infants [of which 5 (20%) had certain HFOs at some point].

To assess whether AORR could predict the development of IESS across different ages, we trained a penalized logistic classifier using a ridge logistic regression model with cubic splines and exhaustive leave-2-subjects-out cross-validation. For the IESS group, only the 19 EEGs recorded before IESS diagnosis (score 1 or 2) were included in the prediction analysis. In addition, only EEGs recorded before 6 months of age were included (patients *n* = 32, EEGs *n* = 76), as after this age there was only one sample from the IESS group (Fig. 2).

In internal validation, the model demonstrated good overall discrimination [AUC 0.802 (95% CI 0.668, 0.936)] and acceptable overall prediction error (Brier score = 0.122). Sensitivity was 0.556, specificity 0.983 and accuracy 0.882. Misclassification details (*n* = 9; 1 false positive, 8 false negatives) and model stability are further described in the Supplementary Material (chapter 2.3).

The final model was fitted in all data, and the predicted risk surface is shown in Fig. 4. The age-specific AORR cutoff for an EEG predicting IESS was lower at younger ages, suggesting that even moderately elevated AORR is clinically significant in the youngest infants, whereas higher AORR values were required to indicate a similar risk in older infants.

**Figure 4.**
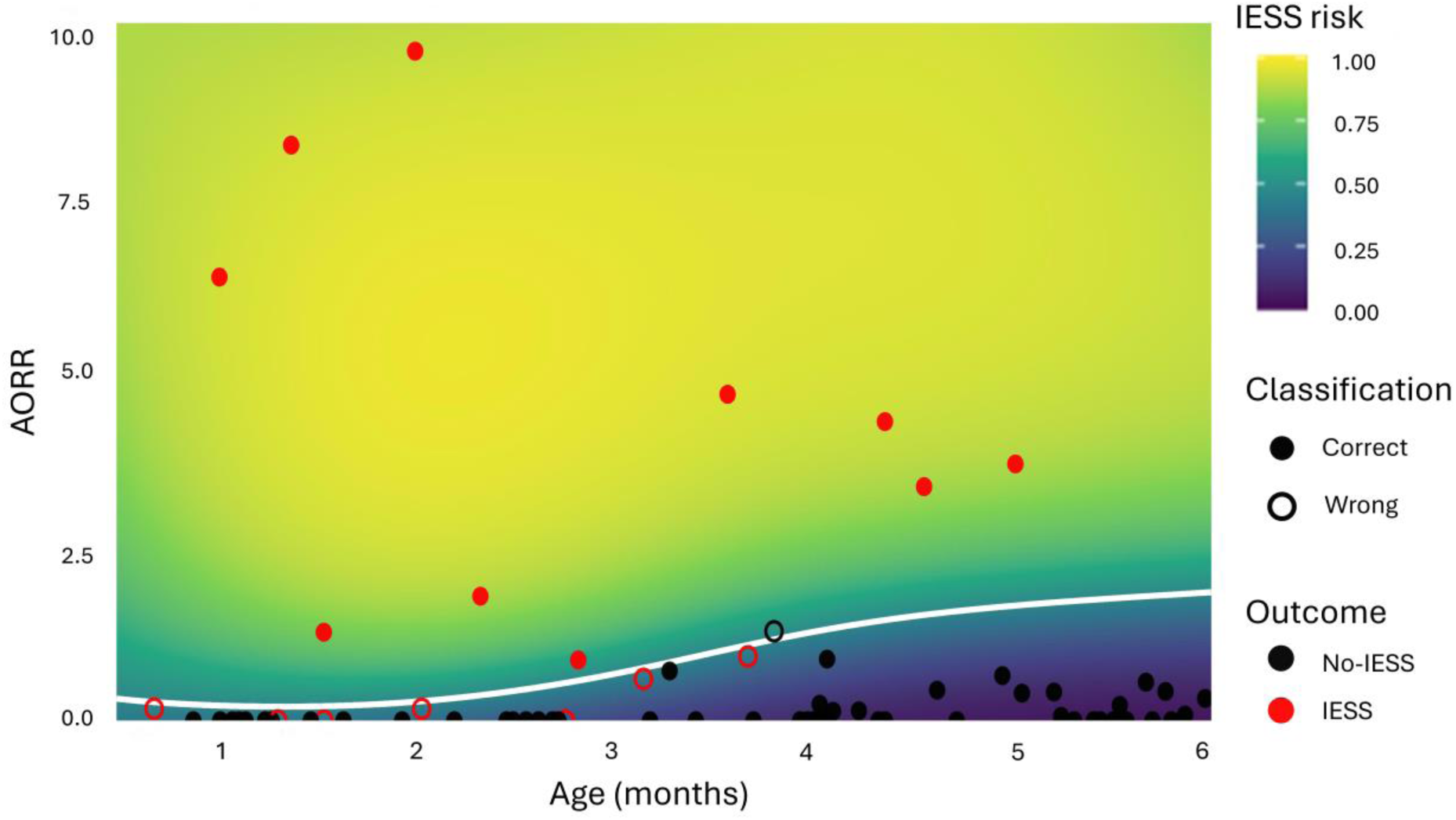
Heatmap of predicted IESS risk. The heatmap represents the predicted IESS (infantile epileptic spasms syndrome) risk from the final spline-based ridge logistic regression model fitted on the full dataset. The colour scale reflects the predicted probability of IESS as a function of age and average overall ripple rate (AORR). The white curve represents the model-based decision boundary (age-specific AORR cutoff), defined as the combinations of age and AORR for which the predicted probability equals the decision threshold. This threshold (0.461) was chosen so that the classifier achieved at least 95% specificity while simultaneously keeping sensitivity as high as possible. Age-specific AORR cutoffs at this threshold were 0.22, 0.27, 0.67, 1.27, 1.64, and 1.84 at 1, 2, 3, 4, 5 and 6 months, respectively. Overlaid points show cross-validation predictions: IESS samples are shown in red and no-IESS samples in black; correctly classified EEG samples are displayed as solid circles, and misclassified samples as open circles with a coloured outline corresponding to the diagnostic class.

Secondarily, we repeated the above analysis using the automatically detected HFOs (80–140 Hz). The findings were qualitatively in the same direction as in the primary analysis; however, the model based on automatically detected HFOs showed somewhat lower performance [AUC 0.724 (95% CI 0.582, 0.866), sensitivity 0.333, specificity 0.914, accuracy 0.776]. See the Supplementary Material (chapter 2.3.2) for further details.

### Interictal epileptiform discharges

The IED criteria were assessed using all eligible 168 EEG recordings, including those excluded from HFO analysis due to low sampling rate or artifacts. The score 2 IED criteria were fulfilled prior to IESS diagnosis in 10/11 infants (one infant had a score 1 EEG at 1 month and the second EEG at 4.5 months was already diagnostic of IESS, score 3), and in additional two infants who did not develop IESS. In these two infants, the IED criteria were no longer met in later EEGs (Fig. 5).

**Figure 5.**
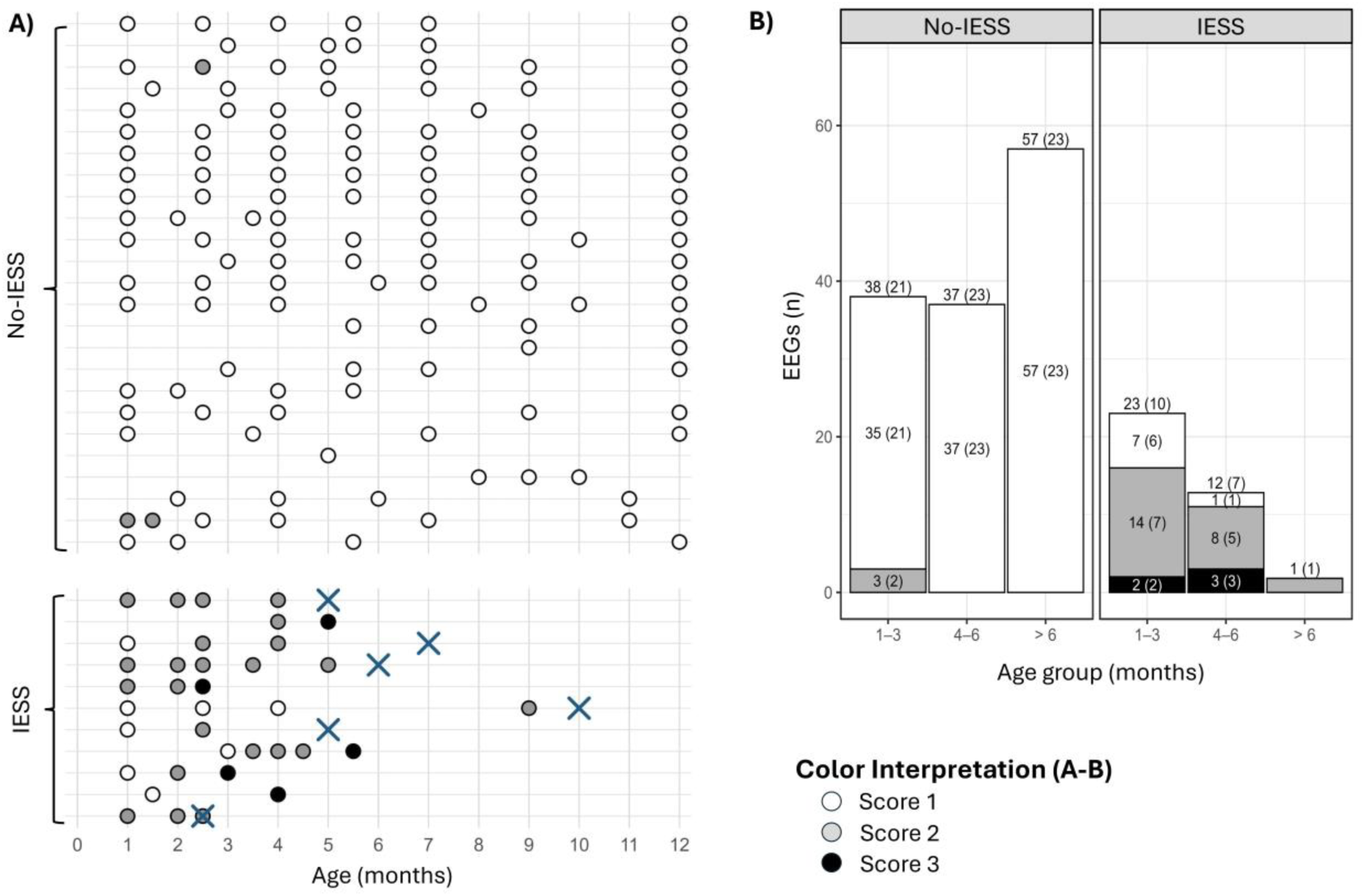
**EEG IED** (interictal epileptiform discharges) **score by IESS** (infantile epileptic spasms syndrome) **status at patient (A) and group (B) levels.** IED score is represented with identical colour coding in both sections A and B. Score 1 (white): No significant epileptiform or seizure discharge findings; Score 2 (grey): Recurrent epileptiform activity; Score 3 (black): EEG diagnostic of IESS. **(A)** All 168 EEGs on a timeline at patient-level. Not all EEGs were suitable for HFO analysis. Each row represents a single patient. Blue X marks the time of IESS diagnosis if the diagnostic IED score 3 EEG was not available due to exclusion of EEGs recorded after vigabatrin initiation (n = 5) or clinical diagnosis of IESS (n = 1). For visualization, ages were rounded to the nearest full month. Some infants underwent additional EEG control recordings, resulting in data points with an accuracy of 0.5 months. **(B)** Distribution of IED scores across age groups, stratified by IESS diagnosis. Because most patients had multiple EEG recordings analysed during the follow-up period, the data were grouped by age for visualization purposes. Bars represent the number of EEG recordings, with segment labels indicating counts (unique patients in parentheses, as a single patient may contribute multiple recordings within the age group).

Fulfilment of score 2 IED criteria strongly predicted the development of IESS during the first six months of life (patients *n* = 34, EEGs *n* = 105). In a ridge-penalized logistic regression with a spline term for age, the model showed good discrimination during internal validation [AUC 0.826 (95% CI 0.720, 0.932), sensitivity 0.733, specificity 0.960, accuracy 0.895, Brier score 0.109]. The model produced 11 (10.5%) misclassifications (Fig. 6A, the Supplementary Material chapter 2.3.1). The result remained essentially the same when including only the 76 EEGs that were also analysed for HFOs (Supplementary Fig. S7).

**Figure 6.**
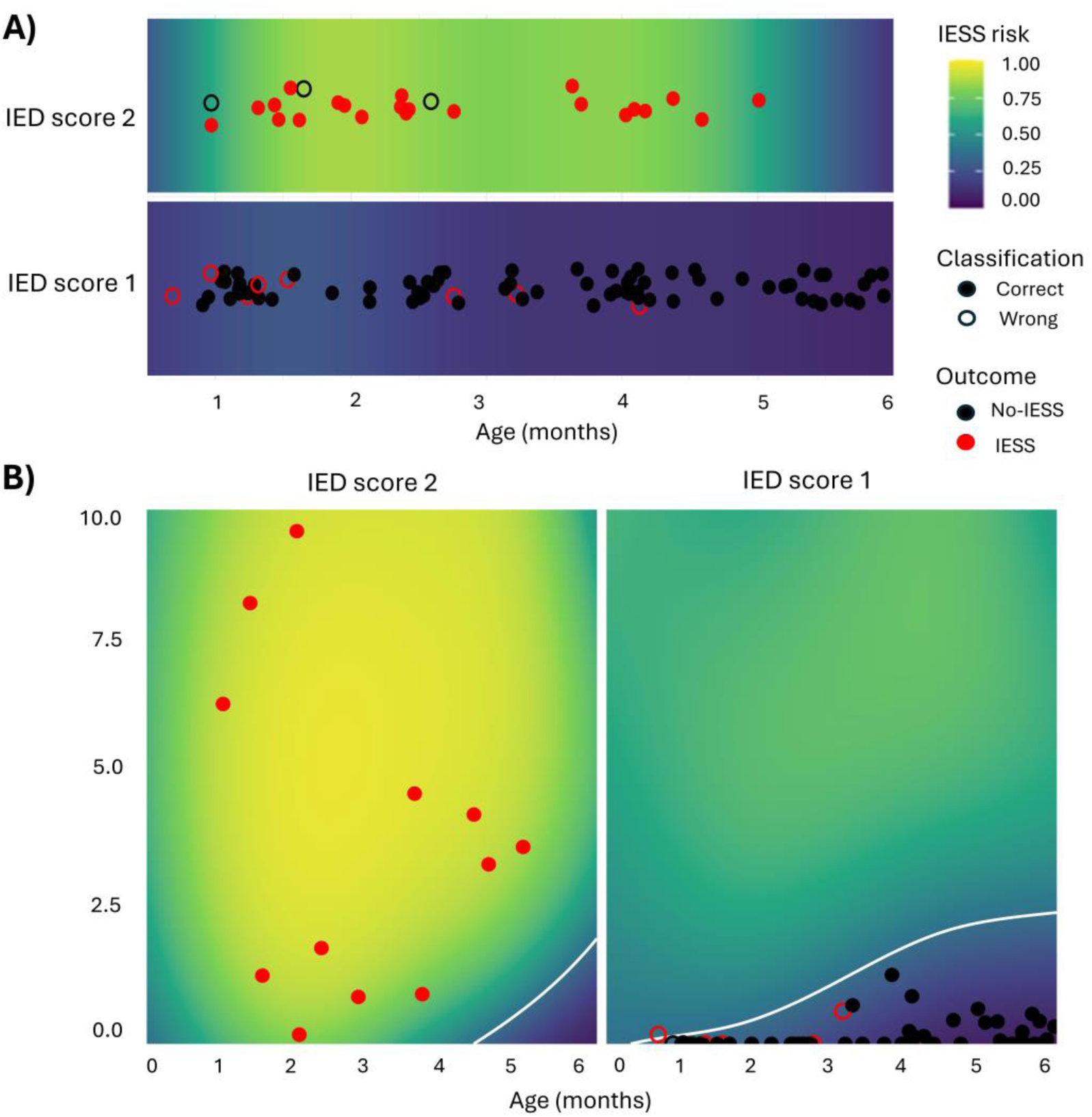
**Logistic regression models for prediction of IESS** (infantile epileptic spasms syndrome). **(A)** Predicted probability of IESS from a ridge-penalized logistic regression model with age and binary IED (interictal epileptiform discharge) score as predictors. Age and its interaction with IED score are modelled using a natural cubic spline. The background heatmap shows the model-based IESS risk as a function of age (x-axis) for EEGs with IED scores 1 and 2 (separate horizontal bands). Overlaid points show internal cross-validation predictions at a clinically motivated probability threshold (0.366) derived from the out-of-fold ROC analysis (specificity ≥95%, with sensitivity maximized under this constraint). IESS samples are shown in red and no-IESS samples in black; correctly classified EEG samples are displayed as solid circles and misclassified samples as open circles with a coloured outline corresponding to the true class. **(B)** The heatmaps represent the predicted IESS risk from the final spline-based ridge logistic regression model fitted on the full dataset, separately for IED score 1 and IED score 2 EEGs. The colour scale reflects the predicted probability of IESS as a function of age and average overall ripple rate (AORR). The white curve represents the model-based decision boundary (age-specific AORR cutoff). The decision threshold (0.366) was chosen so that the classifier achieved at least 95% specificity while simultaneously keeping sensitivity as high as possible. Correctly classified EEG samples are displayed as solid circles and misclassified samples as open circles with a coloured outline corresponding to the true class.

### Comparison of HFOs and IEDs as predictors for IESS

In general, HFOs were detected in most infants fulfilling the IED criteria for Score 2 (9/10 IESS; 1/2 no-IESS), and HFOs appeared at the same time as the IED criteria were fulfilled (6/9 IESS, 0/1 no-IESS). In two cases that developed IESS, HFOs were, however, detected earlier. In four cases, where HFOs were either not detected at all (one IESS, one no-IESS) or were detected later than the IED criteria were met (one IESS, one no-IESS), the relevant EEG recordings could not be evaluated for HFOs due to artefacts or low sampling frequency. In contrast, 17/25 (68%) infants exhibited HFOs without fulfilling the EEG IED criteria at any point, and none of these infants developed IESS. The median time from fulfilment of score 2 IED criteria to IESS diagnosis was 43 days (range 0–134; in one infant with 0 days delay, the IED score 2 spike criteria were only met when the EEG was already diagnostic of IESS i.e. score 3), and from HFO occurrence to IESS diagnosis 58 days (range 0–134).

Finally, we performed a multivariable penalized logistic regression including IED score, AORR and age (patients *n* = 32, EEG *n* = 76), using the same penalized modelling framework as the two-predictor models (Fig. 6B). Pre-analytic assessments indicated no collinearity to a degree that would prevent joint modelling. In internal validation, overall discrimination was good [AUC 0.822 (95% CI 0.684, 0.960), sensitivity 0.667, specificity 0.960, accuracy 0.895, Brier score 0.099]. The joint model produced 6 misclassifications (false negatives) that were identical to the misclassified samples in the IED score and age only model when using the same 76 EEGs (see the Supplementary Material). This multivariable model (Fig. 6B) suggests that IED score 2 is the strongest single warning signal for forthcoming IESS, and in this small sample AORR did not add prediction value of the model.

## Discussion

Our longitudinal cohort study in infants with a large, acquired brain injury demonstrates a clear difference in scalp HFOs between infants who developed IESS and those who did not. First, HFOs were present already in the early EEGs recorded at <4 months of age in most infants developing IESS, unlike those who did not. Second, HFO rate increased with age at a much steeper pace in the IESS group than in the no-IESS group. Third, although HFO rate accurately predicted the development of IESS, the presence of recurrent IEDs (score 2) was as robust as a predictor and is more easily applicable into clinical practice.

### HFOs during epileptogenesis

Intracranial recordings from both animals and humans show that pathological HFOs are indicators of epileptogenic tissue^42–47^ and possibly reflect epileptogenicity even better than interictal spikes.^48^ Animal models also suggest that pathological HFOs are linked to epileptogenesis.^20–22,49,50^ Mechanistically, pathological HFOs are thought to reflect synchronized firing of abnormally bursting principal cells in small discrete and pathologically interconnected neuronal clusters, which, in turn, would initiate epileptogenesis via a kindling effect.^43,51,52^ In rats, only individuals that developed spontaneous seizures after kainite-induced status epilepticus exhibited HFOs.^20^ Likewise, in a traumatic brain injury rat model, pathological HFOs occurred among all animals with later seizures,^21^ were the first EEG abnormality to be observed,^22^ and fast-ripple rates were also higher in animals that later developed epilepsy than in those that did not.^50^

Our results corroborate these findings from animal studies, as we showed both a clear distinction in HFO occurrence and rates between the IESS and no-IESS groups, and a steep increase in HFO rates towards the IESS diagnosis. Considering that all included infants had a large, acquired brain injury, the earlier occurrence and increased HFO rates in the IESS group likely reflect epileptogenesis and are not explained by the brain injury itself. This conclusion is also supported by our previous study in neonates showing that neonatal HFOs were associated with established or forthcoming neonatal or infantile onset developmental and epileptic encephalopathy (DEE),^15^ whereas healthy neonates did not express certain HFOs.^15,53^ While we suggest that early-occurring HFOs are likely pathological, the interpretation of HFOs becomes increasingly complex with growing age, and at least later in childhood, the physiological HFOs appear to exhibit complex developmental dynamics.^54^ Studies indicate that physiological HFOs increase during the first year of life,^18,55,56^ and indeed, the HFO rate also slowly increased in our no-IESS group. Taken together, the mere existence of HFOs is likely not informative as to the risk of epilepsy later in infancy, whereas HFO rate may still distinguish those at risk as it was clearly higher in the IESS group even after the first months of age.

An optimal biomarker for epileptogenesis should be specific for epilepsy, translatable (i.e. applicable both in therapy development among animals and in clinical antiepileptogenesis trials among humans), non-invasive, stable in its expression and economically feasible.^57,58^ Based on previous literature and our current results, we suggest that the scalp HFOs carry high potential to be early biomarkers of epileptogenesis, but further research and validation in other aetiologies and larger cohorts are needed.

### Prediction of IESS development

Studies investigating how EEG evolves towards IESS are scarce. In a retrospective study of children who later developed IESS, EEGs during a clinically asymptomatic phase (i.e. weeks to months prior to the onset of spasms) showed (multi)focal spike-slow-wave activity not exceeding 50% of the NREM EEG recording time, with at most mild background abnormalities.^11^ If the EEG normalized from mild background abnormality, IESS did not develop, and if modified hypsarrhythmia was observed, it progressed towards IESS in the absence of treatment.^11^ In another study of preterm infants with periventricular leukomalacia (PVL), bilateral parieto-occipital multi-spike-slow-wave complexes at 3 months of corrected age, always indicated forthcoming IESS.^59^ Finally, in infants with HIE, once multifocal spikes appear on EEG, the delay to the onset of spasms is short.^9,13^

In the current study, HFO rate accurately predicted the development of IESS. However, given the previous literature and as reliable HFO analysis is time-consuming and poses additional demands on EEG data quality, we critically evaluated whether analysing HFOs adds to the predictive value of standard visual EEG analysis based on IEDs.^9,12^ In accordance with our previous results,^9,12^ the EEG-based IED criteria effectively identified infants later developing IESS. Also, the specificity was high, meaning that a score 1 EEG (i.e. no significant epileptiform or seizure discharge findings) was highly suggestive of no-IESS outcome, especially after age 2–3 months. In a multivariable model, HFOs did not provide additional predictive value to IEDs. However, in two infants developing IESS, HFOs appeared in the EEG earlier than the IED criteria were fulfilled, and in our previous study, HFOs were detected already in the neonatal EEGs of some of the infants later developing IESS.^15^ Hence, early-occurring HFOs might be the earlier indicator of forthcoming IESS and help to identify ongoing epileptogenesis even when the IED score is still normal, but further data are needed to convincingly validate this finding, and to evaluate its possible clinical implication. If prophylactic treatments for IESS were to be established, it would be crucial to correctly identify infants at the highest risk of IESS as early as possible.

### Limitations

Despite the population-based study design, our actual cohort was small. Furthermore, the timing of EEG recordings was heterogenous, and the number of EEGs per infant remained limited especially in the IESS group as the IESS diagnoses were made already during the first few months of life. Despite these limitations, our data showed a very robust association of HFO rate and IED score with development of later IESS.

In scalp EEGs and especially among infants, artefacts due to movement and muscle activity remain a major challenge for reliable HFO evaluation. Although minor muscle artifacts or sporadic movements do not hamper standard clinical EEG review, they compromise HFO analysis, as they contain high frequency components, which obscure the minute HFOs. In our cohort, artefacts resulted in exclusion of 22/148 (15%) otherwise eligible EEGs that could be analysed for IEDs. Thus, the implementation of HFO analyses in routine clinical practice would require additional effort concerning not only the interpretation but also the quality of EEG data, and certain infant-related factors likely remain impossible to control.

Artefacts also contributed to our inability to determine whether a concomitant analysis of IED criteria and HFOs could be useful. In our cohort, there were two infants exhibiting IEDs who never developed IESS, and it is plausible that the rate and occurrence of HFOs could differentiate the highly pathological IEDs from more benign ones.^60^ Unfortunately, the EEGs fulfilling the IED criteria in these two infants were unsuitable for HFO analysis.

Visual HFO scoring with the currently suggested methods^61^ is inevitably biased, as the standard EEG is simultaneously available to the reviewer and the presence of IEDs may favour the interpretation of HFOs as probable or definite. Regarding automated analysis, artifact contamination remains a challenge. We, nevertheless, had a good interrater agreement, and the agreement between visual and automatic detections was high at lower frequencies (80–140 Hz) when artifacts were likely well-controlled, supporting the reliability and objectivity of the results.

Finally, the study setting and the small cohort did not allow us to completely account for possible effects of medication. Physiological HFOs are not affected by ASM,^62^ whereas pathological HFOs were shown to decrease in animal models,^63–65^ while in humans, the results remain inconsistent.^48,66,67^ Vigabatrin, which is commonly used as one of the first-line treatments for IESS, effectively suppresses pathological HFOs, generation of spasms and hypsarrhythmia in animal models,^64^ and hence, due to its likely HFO modulatory and possibly IESS preventive effect, we excluded infants who received vigabatrin and did not develop IESS. After careful consideration, the infants who developed IESS despite vigabatrin treatment (i.e. when there was no prophylactic effect) were included in the study, but only their EEGs prior to vigabatrin initiation were analysed. Hence, in the included infants the vigabatrin treatment did not affect the HFO analyses, but it might have postponed the onset of spasms. We did not consider ASM received for acute symptomatic seizures during the neonatal period as relevant for later epileptogenesis, since the typical medications used (phenobarbital, levetiracetam, midazolam and fosphenytoin) are not common practice in the treatment of IESS and likely carry no prophylactic effect. We also did not consider other medications received during the neonatal period, as they would not be expected to impact the relationship between the EEG findings and the development of IESS, even if they might impact the overall brain injury severity.

## Conclusion

Our data demonstrate that among infants with a large, acquired brain injury, early (<4 months) occurring HFOs are associated with later development of IESS and are not explained by the brain injury itself. The number of HFOs steeply increases towards the onset of spasms, thereby reflecting the progression of epileptogenesis. However, our robust IED scoring criteria also accurately identify the infants at the highest risk of developing IESS and are more easily adaptable to clinical predictive diagnostics than HFOs. Our longitudinal cohort data also inarguably demonstrate that infants with a large, acquired brain injury need frequent follow-up EEGs from an early age to allow prompt diagnosis and possibly even prevention of IESS in the future.

## Data Availability

Due to the strictly confidential nature of clinical patient data, the individual level raw data cannot be made openly available.

## ABBREVIATIONS

AORR: average overall ripple rate
CS: certainty score
CV: cross validation
EEG: electroencephalography
HFO: high-frequency oscillation
HIE: hypoxic ischemic encephalopathy
ICC: intra-class correlation coefficient
IED: interictal epileptiform discharges
IESS: infantile epileptic spasms syndrome
IQR: interquartile range
L2SO: leave-two-subjects-out
LFU: lost to follow-up
MAD: mean absolute deviation
MRI: magnetic resonance imaging
OFF: pit of fold
ORR: overall ripple rate
QS: quiet sleep

