## Supplementary for "High-frequency oscillations and interictal epileptiform discharges predict infantile spasms"

### 1. Supplementary Information on Material and Methods

#### 1.1. Model selection and sensitivity analysis of alternative mixed-effects models

To optimize the within- and between-subject random-effects mixed-effects model, three candidate random-effects structures were evaluated: random intercept-only, random slope-only, and random intercept with random slope. Akaike information criterion (AIC) favored the intercept-only model (AIC = 434, Akaike weight = 0.73), with the intercept+slope model close (ΔAIC = 2, weight = 0.27) but singular (slope variance = 0), indicating that the random slope term did not account for any additional variability. The slope-only model fit the data substantially worse (ΔAIC = 42.5, weight = 0). Prediction error metrics also supported the intercept-only model (Root mean square error [RMSE] = 0.84, Mean absolute error [MAE] = 0.46 vs 1.53/0.76 for slope-only). We therefore selected the random intercept-only structure as the most suitable.

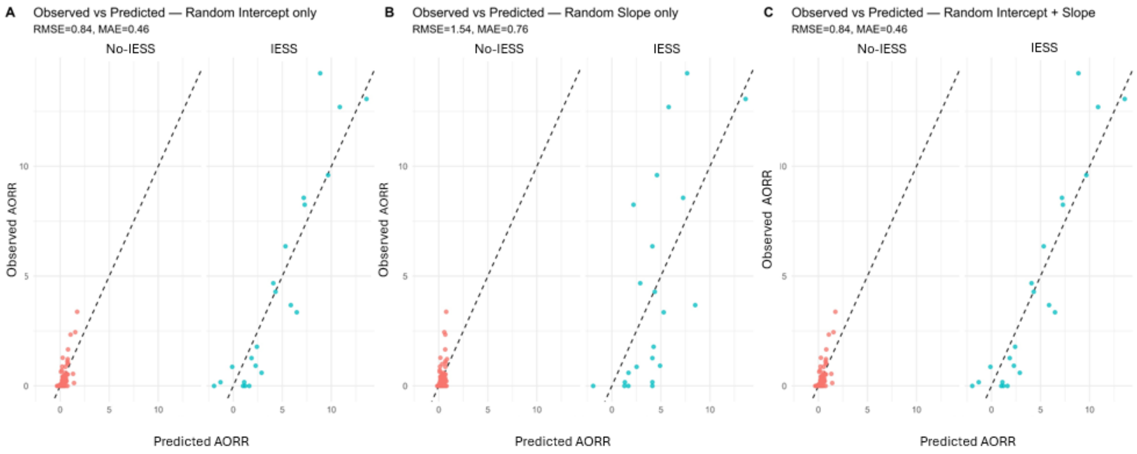


**Supplementary Figure S1 Model-predicted versus observed average overall ripple rate (AORR) values across alternative random-effects structures.** The three structures were (A) random intercept only, (B) random slope only, and (C) combined random intercept and slope. Each point represents one measurement (x = predicted, y = observed); dashed line indicates perfect agreement. Blue dots indicate EEG recordings from patients who developed IESS, whereas red dots indicate recordings from patients who did not develop IESS. The random-intercept model provided the best fit, while the random-slope only model showed the poorest performance. AORR = average overall ripple rate, RMSE = root mean square error, MAE = mean absolute error.

##### 1.2 Ridge-regularized logistic regression model with cubic splines

To examine whether average overall ripple rate (AORR) or the interictal epileptiform discharges (IED) score predicted later development of infantile epileptic spasms syndrome (IESS) across age, we fitted separate L2-penalized logistic regression models using the *glmnet* package in R.[1] Nonlinear effects of the predictor and age were modeled using natural cubic splines with three internal knots placed at the empirical quartiles, and an interaction between the spline-expanded predictor and age terms was included.[2] The ridge penalty (*alpha = 0*), with penalty strength controlled by the tuning parameter, λ, was used to stabilize estimation in the context of a limited sample size.[3,4] The spline-expanded design matrix, including the interaction terms, was standardized by glmnet using the default setting (*standardize = TRUE)*, improving numerical stability and placing the -derived predictor terms on a comparable scale.

Because the model output represents the estimated probability of later IESS for a given EEG recording, each EEG was treated as a separate prediction point. Predictive performance was therefore evaluated at the EEG-sample level. However, to avoid data leakage, all resampling steps, including cross-validation and λ tuning, were performed at the patient level, ensuring that EEGs from the same patient were not split between training and test sets. In addition, during model fitting and λ tuning, inverse-frequency weights of $1 / n_i$ were used to prevent patients with multiple EEG recordings from dominating model estimation, where $n_i$ denotes the number of EEGs available for patient i.

Internal validation relied on exhaustive leave-two-subjects-out cross-validation, in which every possible pair of patients was used once as an independent test set. Because each EEG appears in multiple held-out test sets across the exhaustive resampling procedure, out-of-fold predicted probabilities for the same EEG were averaged before calculating performance metrics.

The tuning parameter λ was selected using the modified bootstrap–cross-validation method of Pavlou et al.[5]. For each leave-two-subjects-out training set, 20 patient-level bootstrap pseudo-datasets with an approximately 25% enlarged pseudo-sample size were generated, five-fold cross-validation was applied within each pseudo-dataset, and the tuned λ was defined as the geometric mean of the resulting *lambda.min* values. This approach was used to stabilize λ selection and reduce variance compared with standard cross-validation in small samples.[5–7]

Model performance and calibration were derived from the aggregated EEG-sample level out-of-fold predictions. Receiver operating characteristic analysis was used to estimate the area under the curve with 95% confidence intervals. For classification, the decision threshold was selected from the aggregated out-of-fold predictions using a clinically motivated criterion. Calibration was evaluated using the calibration intercept, calibration slope, and Brier score.

A final model was refitted on the full dataset using a separately tuned λ obtained via the same Pavlou resampling scheme. The risk heatmap and the age-dependent AORR cut-off curve are presented as final model-derived results.

### 2. Supplementary Information on Results

#### 2.1 Interrater agreement

##### 2.1.1 Interrater agreement on HFO occurrence

Both scorers assigned a certainty score (CS) to every EEG: 1 = definite HFOs, 2 = probable HFOs, 3 = uncertain HFOs, and 4 = no HFOs. Inter-rater agreement between the certainty scores was assessed using Cohen's kappa (Cohen's $\kappa$). When all four original certainty levels (scores 1–4) were maintained, agreement was good: $\kappa$=0.65, $z$=11.2, $p$<0.001. After collapsing scores 1–2 into a single category (certain), while keeping uncertain (score 3) and no HFOs (score 4) as separate categories, overall agreement remained good ($\kappa$=0.67, $z$=10.5, $p$<0.001).

Category-specific (one-vs-rest) analyses indicated good overall reliability of the scoring procedure, with the highest agreement for the certain and no HFOs categories ($\kappa$ = 0.76, z = 8.46, p < 0.001 and $\kappa$ = 0.72 , z = 8.10, p < 0.001, respectively) and somewhat lower—but still moderate to good—agreement for the uncertain category ($\kappa$ = 0.56, z = 6.27, p < 0.001). In addition, disagreements in certainty scores were mainly within neighboring categories (Figure S2).

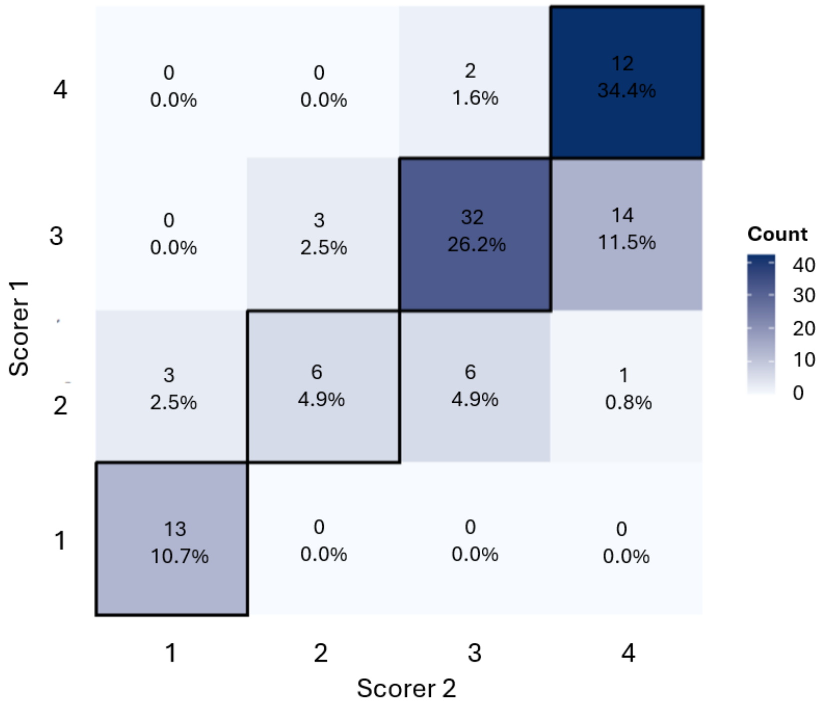

**Figure S2 Heatmap of agreement in HFO certainty scores between the two independent scorers** (rows = Scorer 1, columns = Scorer 2, n = 122). Each cell shows the number of EEGs with that score pair (count; % of total), and shading encodes the percentage (darker = more frequent). The diagonal denotes the exact agreement (n = 93; 76.2%). Certainty score: 1 = definite HFO, 2 = probable HFO, 3 = uncertain, 4 = no HFOs.

##### 2.1.2 Interrater agreement on HFO rate

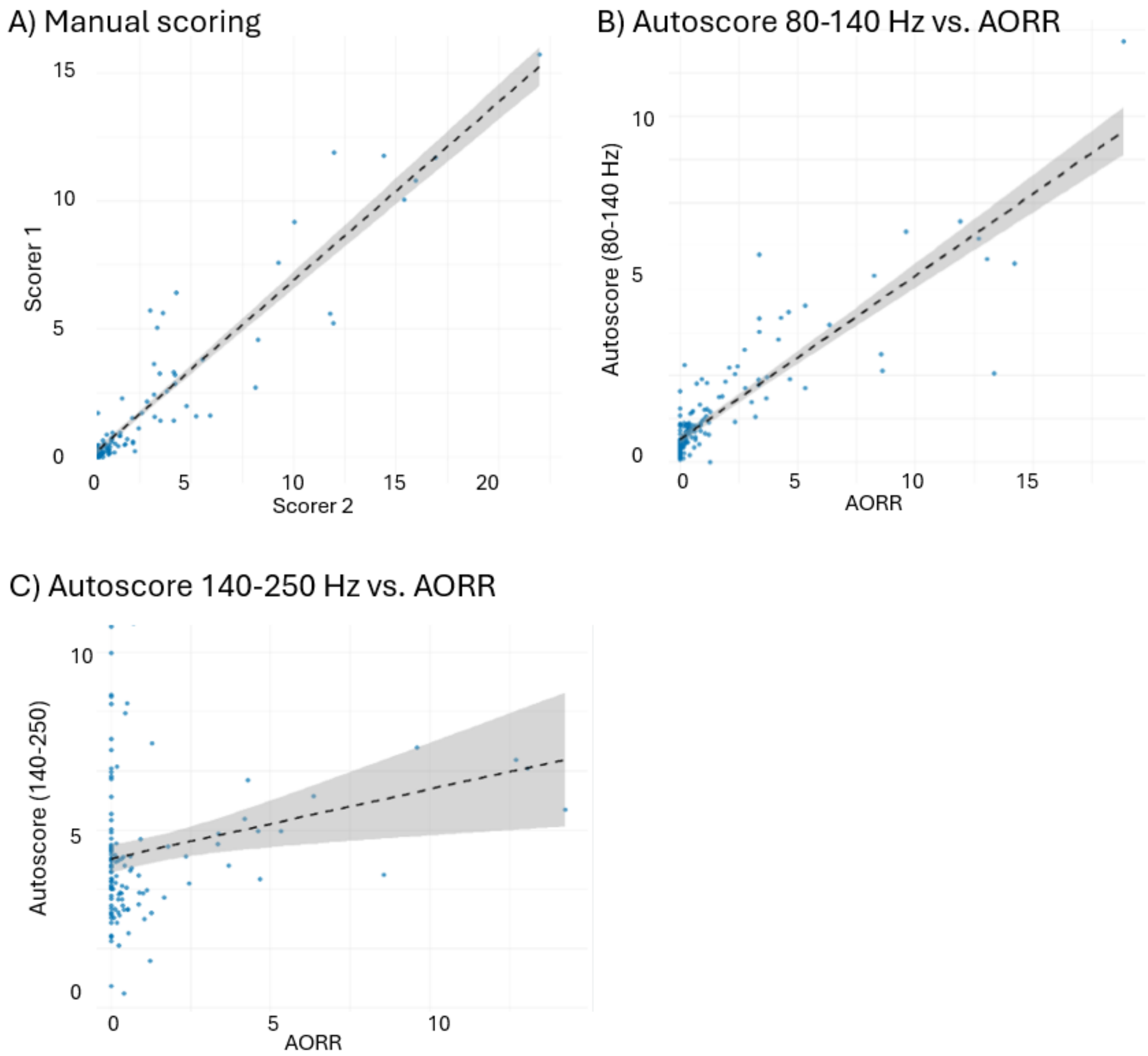


**Figure S3 Scatter plots illustrate the interrater agreement** between A) Scorer 1 and Scorer 2, B) automatic scoring (80-140 Hz) and manual consensus scoring (average overall ripple rate [AORR)], and c) automatic scoring 140-250 Hz and manual consensus scoring (AORR) across all analyzed EEGs. Each point represents a single EEG. The dashed line indicates the best fit based on linear regression, with the shaded area representing the 95% confidence interval. The strong clustering of points along the identity line suggests a high degree of concordance between the two scorers (A) and between the

80-140 Hz automatic detections and AORR (B) across the full range of values. Visual inspection shows that deviations from the line are minimal, and there is no clear evidence of systematic bias. This visual observation is supported by the high intraclass correlation coefficient (ICC), indicating excellent interrater agreement. On the contrary, there was no significant correlation between the 140-250 Hz automatic detections and the AORR (C). ICC in the different scenarios: A) 0.89, 95% CI [0.85–0.91]), $p < 0.001$, B) 0.73, 95% CI [0.66–0.80]), $p<0.001$, C) -0.119, $p=0.907$.

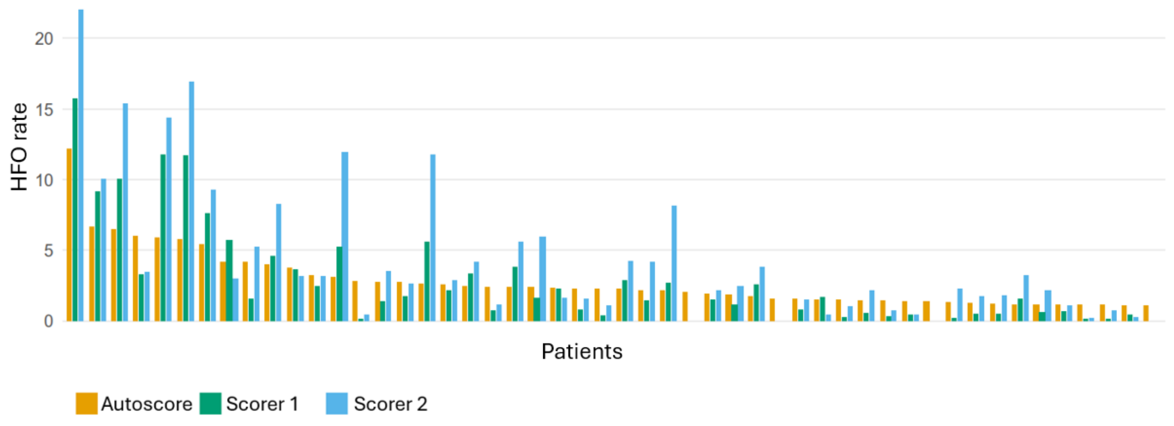


**Figure S4 Comparison of automated detector–derived and expert-scored HFO rates for the 50 highest automated detections.** The orange columns show the 50 highest HFO rates detected by the automatic detector in the 80–140 Hz band, ordered from highest to lowest. For each case, the corresponding HFO rates independently assigned by two expert human scorers are shown side by side in green and blue. Some cases were judged HFO-negative by the human scorers and have no corresponding expert-scored columns.

#### 2.2 Additional results on evolution of HFOs with increasing age (mixed-effects model)

As a secondary analysis, age-related changes in HFO rate were assessed under five different conditions: (a) the rate in the channel with the maximal HFO activity, (b) the rate of HFOs considered certain by the two reviewers, (c) the midline AORR, (d) the rate in the channel with the maximal HFO activity, and (e) the autoscored HFO rate (80-140 Hz) (Table S1).

Focal AORR values were calculated in the same way as the global AORR, but limited to the relevant electrode groups: midline (Fz–Cz, Cz–Pz), left hemisphere (Fp1–F7, F7–T3, T3–T5, T5–O1, Fp1–F3, F3–C3, C3–P3, P3–O1), and right hemisphere (Fp2–F8, F8–T4, T4–T6,

T6–O2, Fp2–F4, F4–C4, C4–P4, P4–O2). AORR calculation procedure is described in detail in our previous publication[8].

Across all these conditions, infants that developed infantile epileptic spasms syndrome (IESS) showed a strong and consistent positive within-person association between age and HFO rate, whereas this association was weaker or, for certain HFOs, absent in children that did not develop IESS. Between-person effects (i.e., differences between children of different mean ages) were small and nonsignificant across all conditions, indicating that the group difference does not arise from age differences between children, but reflects a significantly steeper age-related increase in HFO rate among IESS children compared to those surviving without IESS. The HFO–age relationship difference between the two groups was statistically significant across all conditions. However, the magnitude of this age-related difference varied by analysis type, being smallest for the autoscored condition and largest when using the rate from the channel with maximal HFO rate. These findings are consistent with the results (overall/global AORR) reported in the main article.

| Condition | Group | Within-person slope (β, 95% CI, p) | Between-person effect (β, 95% CI, p) | Group difference in slopes (Δβ, 95% CI, p) | Random effects (SD intercept / residual) | Intra-class correlation coefficient |
|---|---|---|---|---|---|---|
| **Channel with maximal HFO rate** | IESS | 8.60 (7.33–9.87), <0.001 | 0.75 (−2.53–4.03), 0.647 | +8.36 (7.06–9.65), <0.001 | 7.18 / 3.82 | 0.78 |
| | No-IESS | 0.25 (0.02–0.48), 0.035 | 0.24 (−1.52–2.00), 0.787 | | | |
| **Certain HFOs only** | IESS | 2.43 (2.11–2.75), <0.001 | 0.39 (−0.32–1.11), 0.272 | +2.41 (2.08–2.74), <0.001 | 1.51 / 0.97 | 0.71 |
| | No-IESS | 0.02 (−0.04–0.08), 0.495 | 0.05 (−0.33–0.43), 0.797 | | | |
| **Midline ORR** | IESS | 2.43 (2.11–2.75), <0.001 | 0.39 (−0.32–1.11), 0.272 | +2.41 (2.08–2.74), <0.001 | 1.51 / 0.97 | 0.71 |
| | No-IESS | 0.02 (−0.04–0.08), 0.495 | 0.05 (−0.33–0.43), 0.797 | | | |
| **Hemisphere with higher ORR** | IESS | 2.50 (2.17–2.83), <0.001 | 0.67 (−0.25–1.58), 0.093 | +2.45 (2.18–2.72), <0.001 | 1.42 / 0.83 | 0.67 |
| | No-IESS | 0.05 (0.00–0.11), 0.038 | 0.04 (−0.31–0.39), 0.823 | | | |
| **Autoscored rate (80-140 Hz)** | IESS | 0.97 (0.79–1.16), <0.001 | 0.15 (−0.29–0.60), 0.482 | +0.91 (073–1.10), <0.001 | 0.96 / 0.54 | 0.76 |
| | No-IESS | 0.06 (0.03–0.09), <0.001 | 0.16 (−0.08–0.40), 0.177 | | | |

**Table S1 Results of mixed-effects models assessing age-related change in HFO rate across different conditions.**

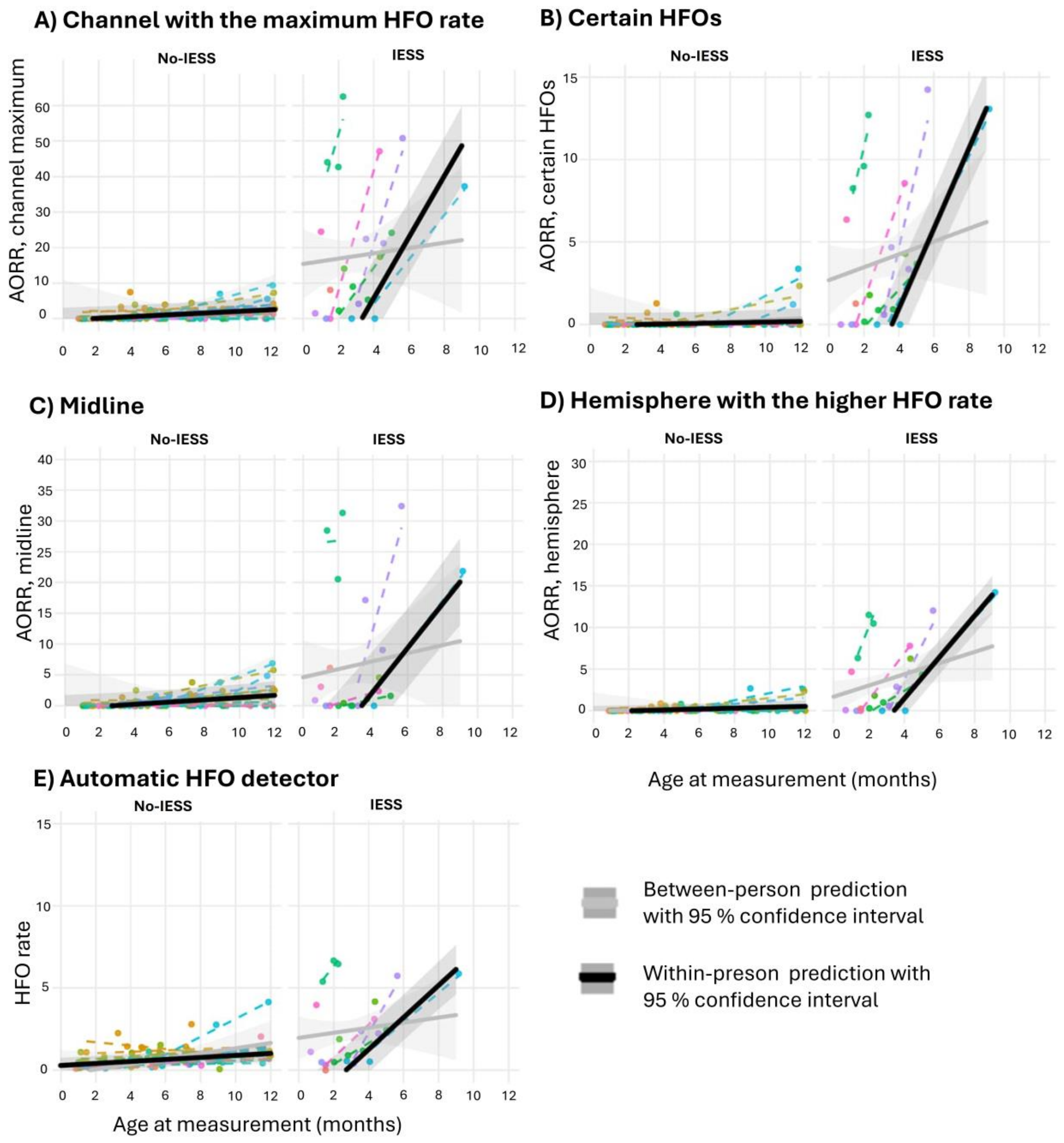


**Figure S5 Age-related HFO rate trajectories in no-IESS and IESS groups across different scenarios.** Mixed-effects model with random intercepts for age and HFO rate in the no-IESS (left panel) and IESS (right panel) groups, together with observed individual patient-level HFO rate trajectories. Results are represented for different scenarios A-E (A = channel with maximum HFO rate, B = certain HFOs, C = average overall ripple rate (AORR) from midline channels, D = AORR from a hemisphere with the highest HFO rate, E = Autoscored rate [80–140 Hz]). Dots show observed AORR values, while dashed colored lines represent raw individual linear fits (for patients with ≥2 measurements). The solid black line represents the population-level fixed-effects prediction at the within-person level with its 95% confidence interval (effect of age change within the same patient). The solid gray line represents the corresponding prediction at the between-person level with 95% confidence interval (effect of differences in average age between patients). In both groups, AORR increases with age within patients in all scenarios, with a markedly steeper slope estimated in the IESS group.

#### 2.3 Additional results on prognostic value of AORR and IED score in IESS (Ridge logistic regression)

##### 2.3.1 Patient-level classification performance (in the main models)

Patient-level classification performance of the main models (chapters 3.2 - 3.3 in the main article) is summarized in Table S2 separately for IESS and no-IESS patients.

In the AORR+age model (*n*=76), there were 10 misclassifications, of which 2 were false positive and 8 false negative. False negatives occurred at low AORR values (≤1) or at AORR = 0 during the first 1–4 months of life. In addition, two no-IESS patients were misclassified at ages 4 and 3 months.

In the IED score+age model (all samples, *n*=105), there were 11 misclassifications (3 false positive, 8 false negative). Eight misclassifications occurred among IED score 1 EEGs recorded between 0.67 and 4.07 months of age (all false negative). The remaining three occurred among IED score 2 EEGs recorded between 0.98 and 2.66 months, all of which were false positive; Six of these EEGs (all false negative) were available for HFO analysis and were also misclassified in the AORR+age model. In contrast, two EEGs that were misclassified as false negative (due to low AORR values < 1 between 1–4 months of age, patient 27) and one EEG that was misclassified as a false positive (AORR 1.28 at 4 months, patient 2) in the AORR+age model were correctly classified by the IED score+age model.

In the multivariable model (AORR+IED score+age, *n*=76), the six EEGs that were misclassified using the IED score+age model and were analysed for HFOs, were still misclassified as false negative.

| No-IESS | | | | | | |
|---|---|---|---|---|---|---|
| | Samples (n) | | Accuracy | | | Misclassified EEGs |
| **Patient** | **AORR** | **IED score** | **AORR** | **IED score** | **Multivar.** | |
| **1** | 2 | 4 | 1 | 1 | 1 | |
| **2** | 3 | 4 | 0.67 | 0.75 | 1 | At 3.8M AORR 1.28 and IEDS1 (AORR model), at 2.7M IEDS2 (IED model, no HFO data) |
| **3** | 3 | 3 | 1 | 1 | 1 | |
| **4** | 4 | 4 | 1 | 1 | 1 | |
| **5** | 3 | 4 | 1 | 1 | 1 | |
| **6** | 3 | 4 | 1 | 1 | 1 | |
| **7** | 4 | 4 | 1 | 1 | 1 | |
| **8** | 4 | 4 | 1 | 1 | 1 | |
| **9** | 4 | 4 | 1 | 1 | 1 | |
| **10** | 4 | 4 | 1 | 1 | 1 | |
| **11** | 3 | 3 | 1 | 1 | 1 | |
| **12** | 3 | 4 | 1 | 1 | 1 | |
| **13** | 3 | 3 | 1 | 1 | 1 | |
| **14** | 1 | 1 | 1 | 1 | 1 | |
| **15** | 2 | 2 | 1 | 1 | 1 | |
| **16** | 3 | 4 | 1 | 1 | 1 | |
| **17** | 3 | 3 | 1 | 1 | 1 | |
| **18** | 1 | 2 | 1 | 1 | 1 | |
| **19** | 1 | 1 | 1 | 1 | 1 | |
| **20** | 1 | 3 | 1 | 1 | 1 | |
| **21** | 3 | 3 | 1 | 1 | 1 | |
| **22** | 0 | 4 | NA | 0.5 | NA | At 1.0M and 1.6M IEDS2 (no HFO data) |
| **23** | 0 | 3 | NA | 1 | NA | NA |

| IESS | | | | | | |
|---|---|---|---|---|---|---|
| | Samples (n) | | Accuracy | | | Misclassified EEGs |
| **Patient** | **AORR** | **IED score** | **AORR** | **IED score** | **Multivar.** | |
| **24** | 1 | 4 | 1 | 1 | 1 | |
| **25** | 1 | 1 | 1 | 1 | 1 | |
| **26** | 1 | 3 | 1 | 0.67 | 1 | At 0.9M IEDS1 (no HFO data) |
| **27** | 4 | 5 | 0.5 | 1 | 1 | At 2.0M AORR 0.17 and at 3.7M AORR 0.92 (both IEDS2) |
| **28** | 2 | 2 | 1 | 1 | 1 | |
| **29** | 2 | 3 | 0 | 0 | 0 | At 2.8M and 4.1M AORR 0. At 1.3M IEDS1 (no HFO data). IESS diagnosis at 9.6M |
| **30** | 1 | 2 | 0 | 0.5 | 0 | At 1.3M AORR 0, IEDS1 |
| **31** | 3 | 4 | 0.67 | 0.75 | 0.67 | At 3.2M AORR 0.60, IEDS1 |
| **32** | 1 | 2 | 0 | 0.5 | 0 | At 0.7M AORR 0.17, IEDS1 |
| **33** | 1 | 1 | 0 | 0 | 0 | At 1.5M AORR 0, IEDS1. The 2nd sample was already IEDS3 (excluded). |
| **34** | 1 | 3 | 1 | 1 | 1 | |

**Table S2 Patient-level classification performance.** The table summarizes the number of EEGs included in the patient-level models using only recordings obtained before 6 months of age: 1) AORR + age, 2) IED score + age, and 3) multivariable model (AORR + IED score + age). Accuracy and additional information on misclassified EEGs are also shown. Two patients with all their EEGs recorded after 6 months of age are not shown in the table. In addition, one misclassified IESS patient (patient 29) had IED score 2 and HFO-positive EEGs recorded only after 6 months of age. Another misclassified IESS patient (patient 33) had the second recording at 4 months, that was already diagnostic for IESS (IED score 3), and this EEG was excluded from the analysis. AORR = average overall ripple rate, IEDS = interictal epileptiform discharges score, IESS = infantile epileptic spasms syndrome, M = months.

##### 2.3.2 Supplementary model with automatically detected HFO rate

In addition, to assess the consistency of the results of ridge logistic regression with cubic splines between automatic and manual scoring, we created a new model using age and automatically detected HFO rate (within the 80–140 Hz frequency band) as predictors. Based on the visual assessment of the data and some preliminary testing, the 95% specificity threshold was not reasonable, so we set the threshold to be at the same level (0.461) as in the original AORR model based on manual HFO scoring. Results were parallel to the original AORR+age model, but in internal validation, the performance of the autoscore model was inferior to that of the original model: AUC 0.724 (95% CI 0.582–0.866), sensitivity 0.333, specificity 0.914, and accuracy 0.776. Brier score 0.166. There were 17 misclassifications in total (5 false positives, 12 false negatives).

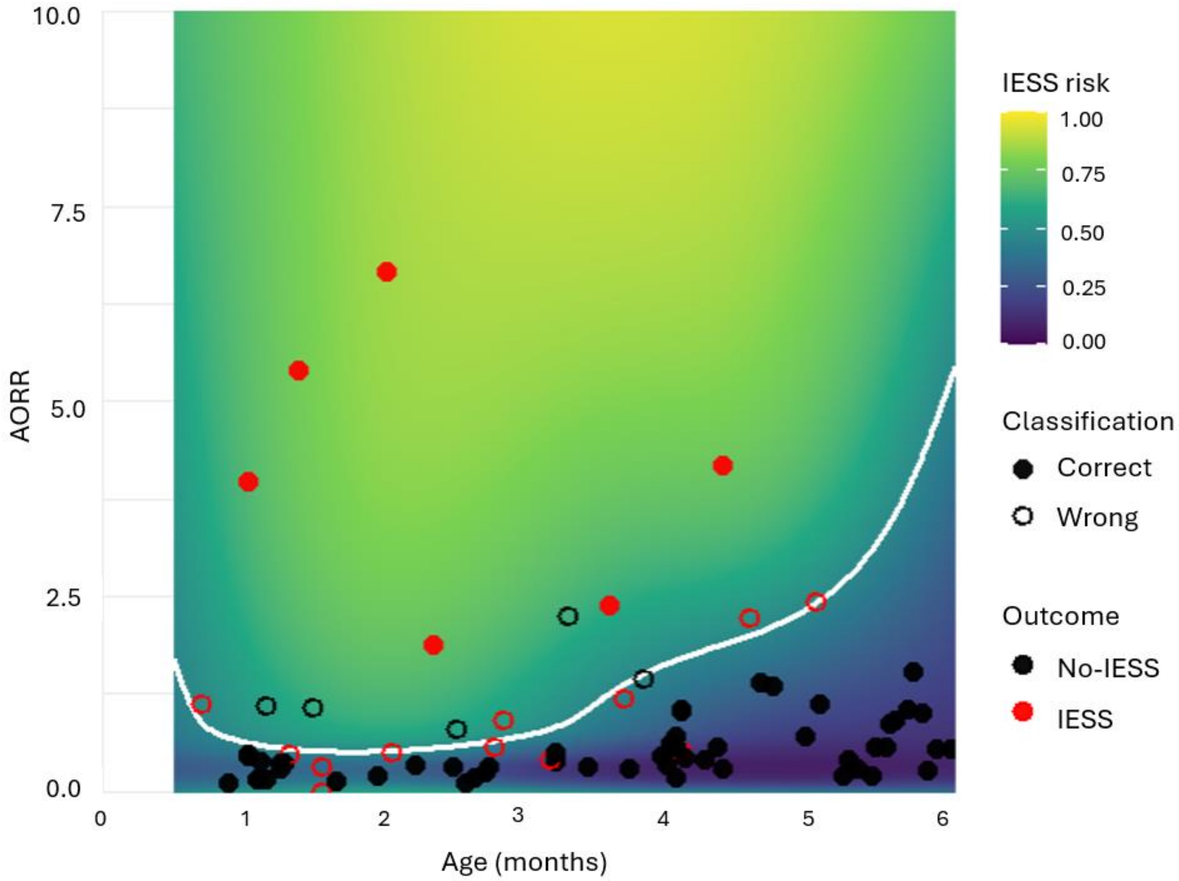


**Figure S6 The heatmap shows the predicted infantile epileptic spasms syndrome (IESS) risk from the final spline-based ridge logistic regression model with automatically detected HFO rate and age as predictors.** The color scale depicts the predicted probability of IESS as a function of age and automatically detected HFO rate. The white curve represents the model-based decision boundary (HFO rate cutoff) at the decision threshold. The threshold was set to be the same as in the manually scored HFO model (0.461). Overlaid points depict cross-validation predictions: EEGs from IESS patients are shown in red and those from no-IESS patients in black; correctly classified EEG samples are displayed as solid circles, and misclassified samples as open circles with a colored outline corresponding to the diagnostic class.

##### 2.3.3 IESS prediction based on IED score: Supplementary model including only EEGs scored for HFOs

Since we had more EEGs available in IED score analysis than in HFO analysis, we assessed the predictive power of IED score, including only those EEGs that were accepted in the HFO analysis.  These models were constructed similarly described in the Supplementary chapter 1.2. The performance was almost identical to the original multivariable model (AUC

0.87 [95% CI 0.694–0.960, specificity 1.00, sensitivity 0.667, accuracy 0.921, Brier score 0.098) and the same 6 individuals were misclassified by both models.

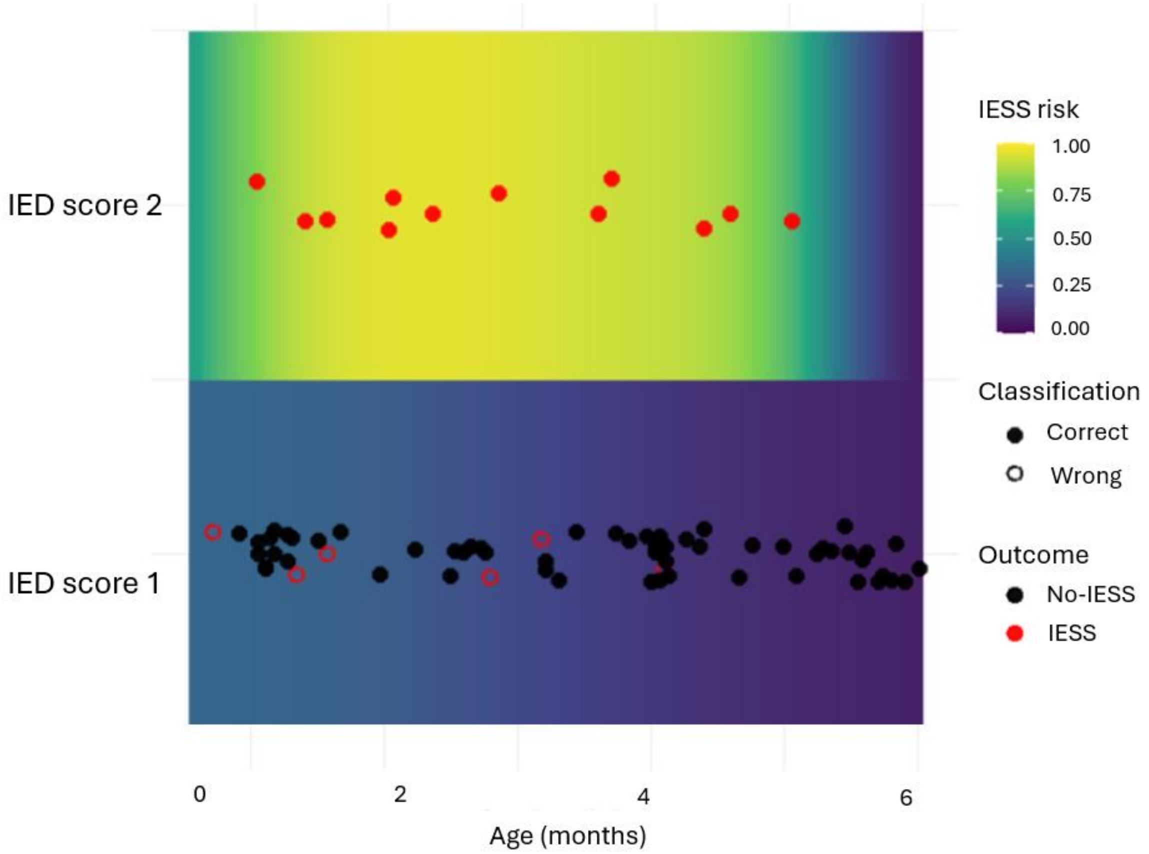


**Figure S7 Predicted probability of infantile epileptic spasms syndrome (IESS) from a ridge-penalized logistic regression model with age and binary IED (interictal epileptiform discharge) score as predictors.** Only EEGs scored for HFOs are included (n=122). The background heatmap shows the model-based IESS risk as a function of age (x-axis) for EEGs with IED score 1 and 2 (separate horizontal bands). Overlaid points show internal cross-validation predictions at a clinically motivated probability threshold (0. 448) derived from the out-of-fold ROC analysis (specificity ≥95%, with sensitivity maximized under this constraint). EEGs from IESS patients are shown in red and those from no-IESS patients in black; correctly classified EEG samples are displayed as solid circles and misclassified samples as open circles with a colored outline corresponding to the true class.

##### 2.3.4 Receiver operating characteristic (ROC) curves

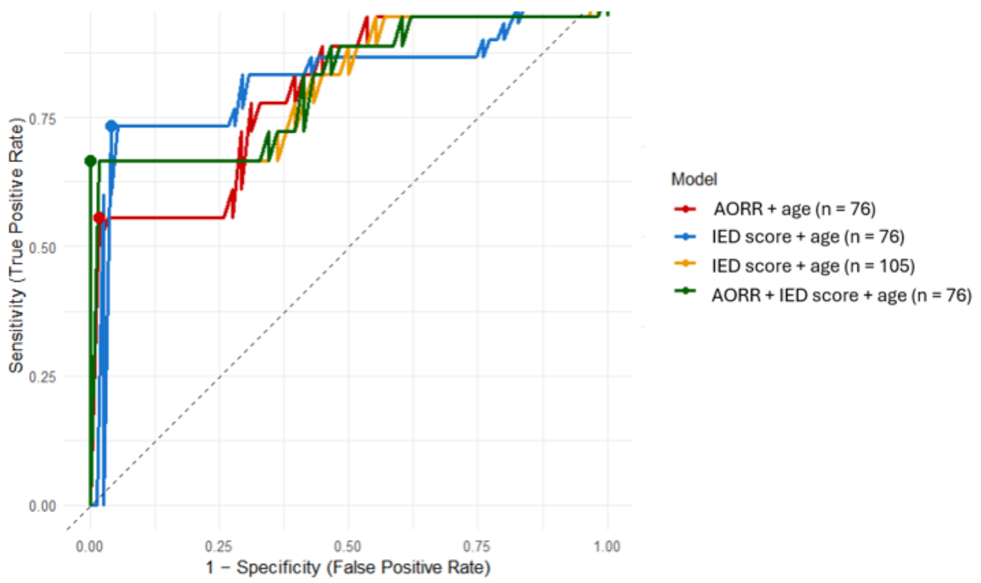


**Figure S8 Receiver operating characteristic (ROC) curve for the ridge–regularized logistic model predicting IESS development.** The curve is based on out-of-fold predictions from patient-grouped exhaustive leave-2-patients-out cross-validation, ensuring no patient-level data leakage. The threshold was clinically motivated (specificity ≥95%, with sensitivity maximized under this constraint). Four different models are concurrently depicted in separate colors: The solid red (average overall ripple rate [AORR] + age), blue (interictal epileptiform discharges [IED] score + age for all data), yellow (IED score + age for EEGs used in HFO scoring), green (AORR + age + IED score) lines.

##### 2.3.5 Model calibration and stability

Across all three ridge-penalized logistic regression models, internal validation indicated acceptable calibration and low prediction instability (Supplementary Table S3). For example, for the model including AORR and age, the mean standard deviation (SD) and mean absolute deviation of predicted probabilities across the 496 exhaustive leave-two-patients-out resamples were 0.025 and 0.016, respectively, indicating low prediction instability. The calibration intercept of –0.428 and slope of 1.18 suggest mild miscalibration, but no evidence of substantial overfitting or prediction instability.

| Model | Calibration intercept | Calibration slope | Prediction stability (mean SD of predicted probabilities) | Prediction stability (mean absolute deviation of predicted probabilities) |
|---|---|---|---|---|
| AORR + age | −0.428 | 1.18 | 0.025 | 0.016 |
| IED score + age | −0.240 | 1.14 | 0.018 | 0.012 |
| IED score + AORR + age | −0.486 | 1.32 | 0.022 | 0.015 |

**Table S3 Ridge-penalized logistic regression calibration and stability metrics.**

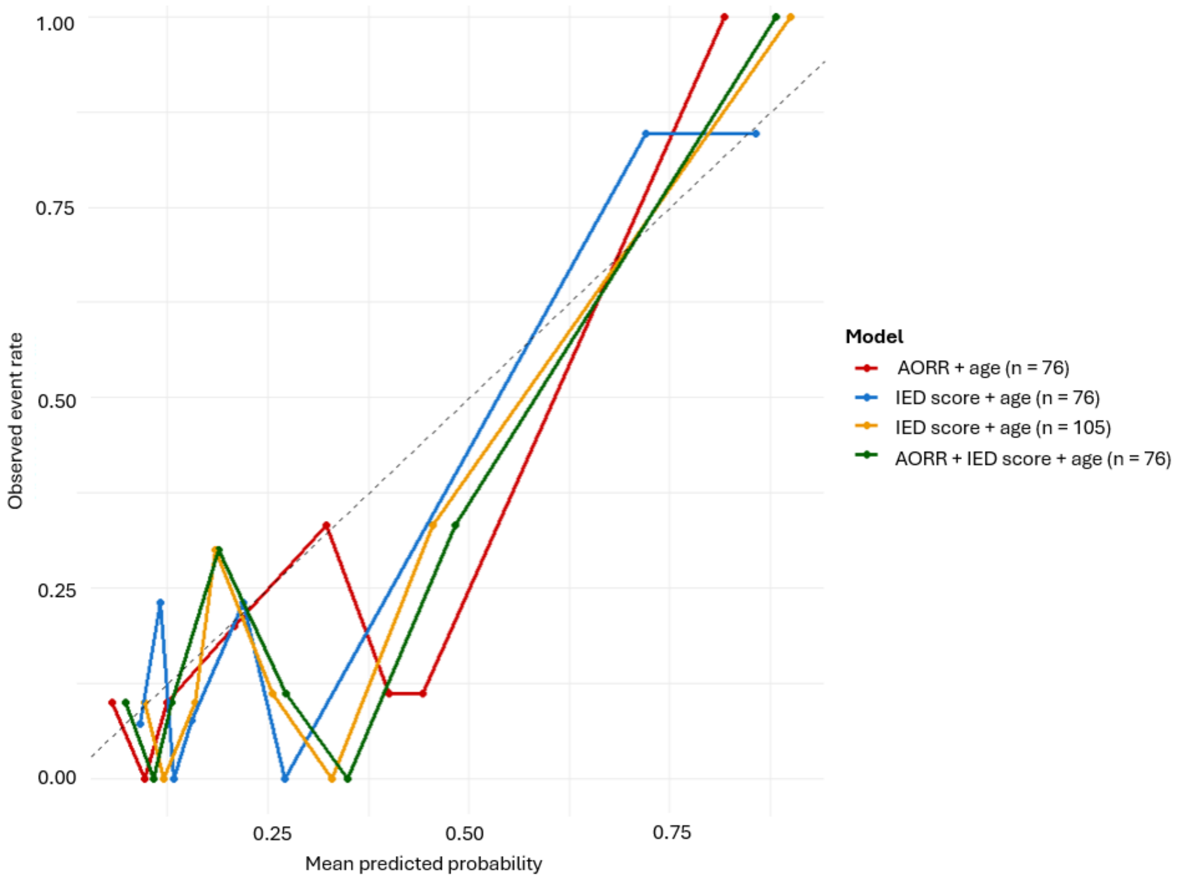


**Figure S9 Calibration curves for the four ridge-penalized logistic regression models based on out-of-fold predictions.** Points show the observed proportion of infantile epileptic spasm syndrome (IESS) cases in groups of patients with similar predicted risk (ten groups ordered from lowest to highest predicted probability). The solid red (average overall ripple rate [AORR] + age), blue (interictal epileptiform discharges [IED] score + age for all data), yellow (IED score + age for EEGs used in HFO scoring), green (joint AORR + age + IED score) lines connect these group-wise estimates, while the grey dashed line indicates perfect calibration. Although there is some deviation at very low and very high predicted risks, all three models show overall acceptable calibration given the limited sample size. Quantitative calibration metrics are provided in Supplementary Table S3.

### References.
